# Clonal haematopoiesis is associated with major adverse cardiovascular events in patients with hypertrophic cardiomyopathy

**DOI:** 10.1101/2024.01.15.24301270

**Authors:** Fernando L. Scolari, Darshan Brahmbhatt, Sagi Abelson, Deacon Lee, Raymond H Kim, Ali Pedarzadeh, Ali Sakhnini, Arnon Adler, Raymond H Chan, John Dick, Harry Rakowski, Filio Billia

**Affiliations:** Peter Munk Cardiac Centre, University Health Network, Toronto, ON, Canada; Ted Rogers Centre for Heart Research, Toronto, ON, Canada; Toronto General Hospital Research Institute, Toronto, ON, Canada; Mount Sinai Hospital, Toronto, ON, Canada; Ontario Institute for Cancer Research, Toronto, ON, Canada; Department of Molecular Genetics, University of Toronto, Toronto, ON, Canada; Institute for Clinical Evaluation Sciences, Toronto ON, Canada; Princess Margaret Cancer Centre, Toronto, ON, Canada; Division of Clinical and Metabolic Genetics, Hospital for Sick Children, Toronto, ON, Canada

## Abstract

**Background & Aims:** The heterogeneous phenotype of hypertrophic cardiomyopathy (HCM) is still not fully understood. Clonal haematopoiesis (CH) is emerging as a cardiovascular risk factor potentially associated with adverse clinical events. The prevalence, phenotype and outcomes related to CH in HCM patients was evaluated.

**Methods:** Patients with HCM and available biospecimens from the Peter Munk Cardiac Centre Cardiovascular Biobank were subjected to targeted sequencing for 35 myeloid genes associated with CH. CH prevalence, clinical characteristics, morphological phenotypes assessed by echocardiogram and cardiac magnetic resonance and outcomes were assessed. All patients were evaluated for a 71-plex cytokines/chemokines, troponin I and B-type natriuretic peptide analysis. Major cardiovascular events (MACE) were defined as appropriate ICD shock, stroke, cardiac arrest, orthotopic heart transplant and death.

**Results:** Among the 799 patients, CH was found in 183 (22.9%) HCM patients with sarcomeric germline mutations. HCM patients with CH were more symptomatic and with a higher burden of fibrosis than those without CH. CH was associated with MACE in those HCM patients with sarcomeric germline mutations [adjusted HR of 3.46 (95% CI 1.25-9.52; p=0.016)], with the highest risk among those that had *DNMT3A, TET2* and *ASXL1* mutations [adjusted HR of 7.23 (95% CI 1.79-29.13) p=0.005]. Several cytokines (IL-1ra, IL-6, IL-17F, TGFa, CCL21, CCL1, CCL8, and CCL17), and troponin I were upregulated in gene-positive HCM patients with CH.

**Conclusions:** These results indicate that CH in patients with HCM is associated with worse clinical outcomes. In the absence of CH, gene-positive patients with HCM have lower rates of MACE.

**KEY POINTS:** *Question:* Does clonal haematopoiesis (CH) affect the phenotype and outcomes among hypertrophic cardiomyopathy (HCM) patients?

*Findings:* Among 799 patients, CH was found in 183 (22.9%) HCM patients. CH was associated with symptoms, a higher burden of fibrosis and major cardiovascular events. Several cytokines and troponin I were upregulated in gene-positive HCM patients with CH.

*Meaning:* CH helps to identify those HCM patients at risk for serious adverse events and unveils a potential inflammatory response mechanism that affects prognosis.

## Introduction

Hypertrophic cardiomyopathy (HCM) is an inherited cardiovascular disorder, affecting 1 in 500 individuals.^1^ However, germline pathogenic or likely pathogenic (P/LP) genetic variants, such as *MYH7* and *MYBPC3* genes, are identified in only 25-40% of patients.^2,3^ A wide heterogeneity and distinct morphologies of left ventricular (LV) hypertrophy are described, even amongst family members carrying similar genetic variants.^1,2,4–7^ Interestingly, interstitial fibrosis is detected in 40-70% of patients and is associated with sudden cardiac death (SCD). Although HCM can be well tolerated, a subset of patients may require advanced treatments such as orthotopic heart transplant (OHT) and LV assist devices.^3,8^

The underlying mechanisms involved in the heterogeneous clinical presentation of HCM patients are not well understood.^3,8^ Clonal haematopoiesis (CH) has emerged as a new risk factor for the development of atherosclerosis, heart failure and cardiogenic shock and is associated with worse outcomes.^9–13^ With aging, haematopoietic stem cells acquire somatic mutations in genes regulating epigenetic DNA modifications and other pathways resulting in clonal expansion. Experiments have shown that CH mutation may promote LV remodelling and fibrosis through increased inflammation.^14–17^ Interestingly, a proteomic study has also shown activation of inflammatory pathways in HCM.^18,19^ The role of CH driving inflammatory processes that may lead to adverse clinical events in individuals with HCM is unknown. Therefore, we sought to evaluate the prevalence of CH in a large HCM cohort and define its association with phenotype and outcomes.

## METHODS

### Patient selection

This is a retrospective study to evaluate the association of CH in a cohort of patients with HCM. We screened 4,400 patients from the HCM clinic of the Toronto General Hospital (Ontario, Canada) from January 2001 to January 2022. We included 799 patients who were > 18 years of age, with a clinical diagnosis of HCM by current guidelines^2,19^, cardiac magnetic resonance imaging (MRI) and available biospecimens from the Peter Munk Cardiac Centre Cardiovascular Biobank. All patients were systematically approached for biobanking. We excluded patients with incomplete records, those that withdraw consent, or low biospecimen quality. This study was approved by the UHN Research Ethics Board (#21-5313) and complied with the Declaration of Helsinki.

### Definitions

HCM was defined as the presence of maximal LV wall thickness (MLVWT) ≥15 mm, ≥13 mm in the presence of a P/LP genetic variant or a family history of HCM, in the absence of other causes. The assessment of P/LP variants was conducted using a previously published strategy by our group^20^. All patients with HCM underwent cardiac MRI with late gadolinium enhancement (LGE) for fibrosis quantification. The LGE was assessed visually and quantified manually as previously validated.^7^ LGE extent was defined as the LGE mass percentage of the total LV mass. We were able to quantify LGE in (84.1%) of patients. In the remaining 127 (15.9%) patients, we only included the qualitative measure of LGE.

### Data collection, outcomes and follow-up

Clinical data were collected from a digital de-identified database. All collected data, including echocardiogram and cardiac MRI, were the closest to the time of biospecimen collection. The following outcomes were collected: appropriate implantable cardioverter defibrillator shock, stroke, cardiac arrest, OHT and death.^21^ We also condensed these as major cardiovascular events (MACE) defined as the first occurrence of any. Only outcomes occurring after sample collection were included. Patients were followed from the time of biospecimen collection to the last visit to our institution. Loss to follow-up was defined as no interaction with the patient at our institution in the preceding one year from chart assessment.

### Clonal haematopoiesis and cytokine assessment

All peripheral blood biospecimens were collected during routine visits to the HCM clinic with consent. For all patients, the biospecimen closest to the cardiac MRI assessment was selected. Briefly, the cytokine analysis was performed using a human cytokine/chemokine 71-plex assay with the Luminex™ 200 system by Eve Technologies Corp. (Alberta, Canada). In addition, the levels of B-type natriuretic peptide and cardiac troponin I were evaluated. For CH genetic analysis, a single molecule molecular inversion probe method targeting 35 myeloid genes related to CH was implemented as previously described^9,22^. To call mutations we used SmMIP-tools^26^. CH was considered in those patients with variants with a VAF ≥2%. Additional details are provided in the Supplementary materials.

### Statistical analysis

The complete statistical methods are shown in the Supplementary materials. Continuous data were evaluated for normality with Shapiro-Wilk test and histogram analysis and presented as mean ± standard deviation or median (interquartile range). Categorical variables were presented as total count (frequency). A Student’s T test or Mann-Whitney test was used to compare continuous variables according to normal distribution. ANOVA and Chi-square test (or Fisher’s exact test) were used for comparing groups. Survival analysis was conducted using the Kaplan-Meier method and differences assessed with the log-rank test. Cox proportional hazards regressions were employed to compare survival rates, adjusting for age as a covariate. A propensity score matching was used to address possible confounders according to CH. As a sensitivity analysis, we also tested those with CH only in the three most common CH genes (*DNMT3A, TET2* and *ASXL1*). We used a statistical significance of 0.05 for all analyses and a two-sided p-value. All analyses were performed using SPSS, version 25.0 (SPSS Inc., NY, USA).

## Results

We included 799 patients in the study and the overall characteristics of the cohort are summarized in (Table 1. Briefly, the mean age of our cohort was 55.7±14.6 years, 547 patients (68.5%) were males, with an age at the time of HCM diagnosis of 47.8±15.8 years. Germline genetic testing was performed on 712 (89.1%) patients, with a P/LP variant identified in 206 (25.8%) patients. P/LP variants were primarily in *MYBPC3* (127, 61.6%) and *MYH7* (53, 25.7%). Mean MLVWT was 17.1±4.2 mm, whereas 170 (21.3%) patients exhibited a LV outflow tract maximal gradient ≥ 50 mmHg. The median time from biospecimen collection to the echocardiogram was 0 (0-16) days and median time from biospecimen to MRI assessment was 2.2 (0.2-5.6) years.

**Table 1.**
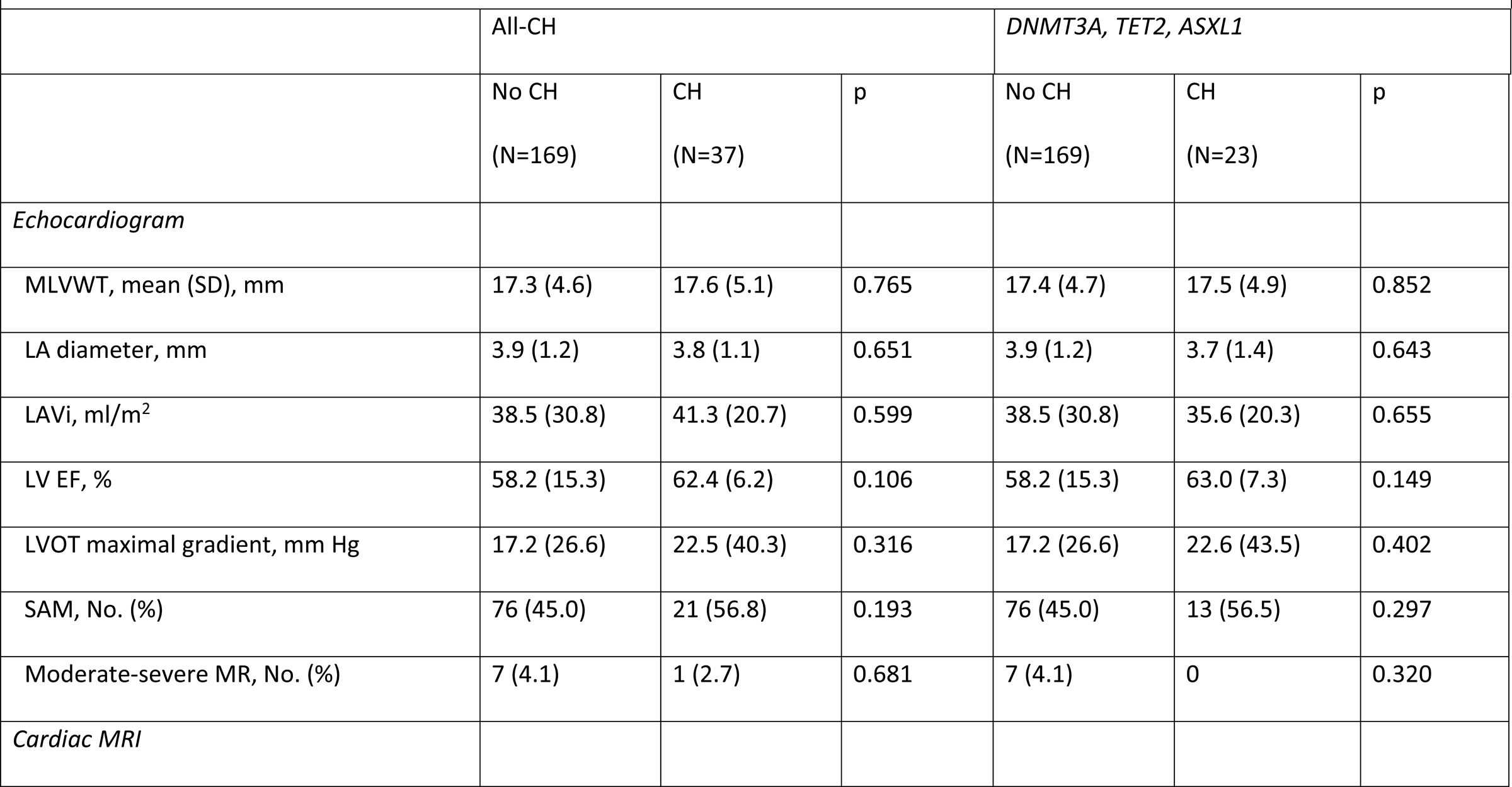
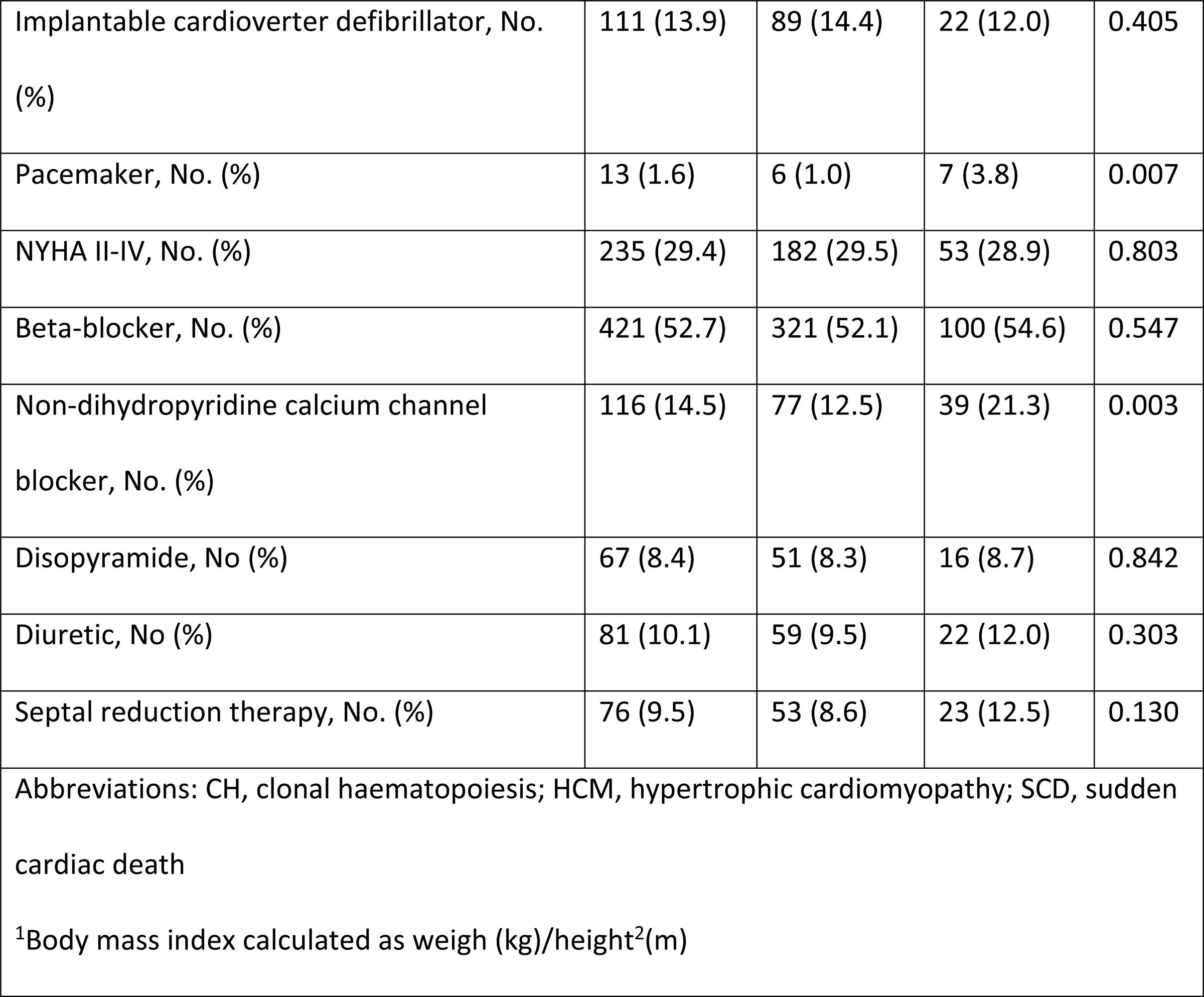
Overall characteristics of the HCM cohort and between those with or without CH.

### Clonal haematopoiesis prevalence and characteristics

All patients had an assessment of CH and all samples passed quality control specifications. CH mutations were observed in 183 (22.9%) patients with a median variant allele (VAF) frequency of 6.7% (2.8-40.8). Of these patients, 136 (17.0%) had somatic mutations in the three most common CH genes: *DNMT3A* in 70 (8.8%), *TET2* in 51 (6.3%), and *ASXL1* in 24 (3.0%), comprising 73.8% of all CH mutations (Figure 1A). The CH-associated gene mutations summary is shown in Supplementary Table 1. Most patients (158, 19.8%) harboured a single mutation and of the 183 patients with CH (Figure 1B), 135 (73.7%) were >50 years old (Figure 1C). In relation to the HCM diagnosis age, the majority, 94 (51.3%), were diagnosed after 50 years old.

**Figure 1.**
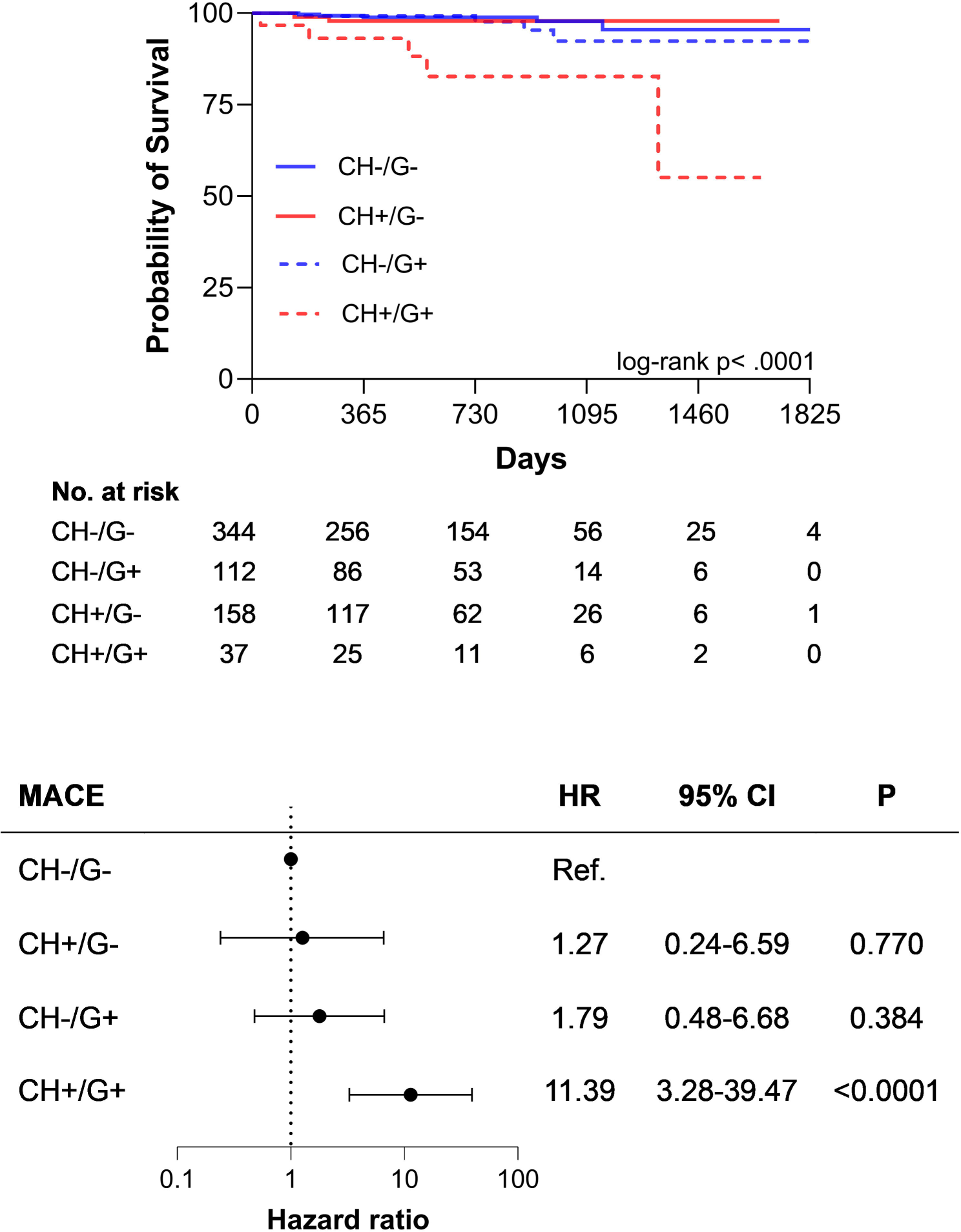
Somatic mutations related to CH in patients with HCM. Panel A shows the number of mutations in the most common affected genes in the cohort.. Panel B shows the number of mutations per patients in those with clonal haematopoiesis .CH. Panel C shows the number of patients with CH according to the decade of the assessment. (CH, clonal haematopoiesis ; HCM, hypertrophic cardiomyopathy).

Overall, clinical characteristics of CH patients were similar to those without CH ((Table 1). There was no statistical difference regarding age, age at HCM diagnosis, and comorbidities according to CH status. Both groups underwent genetic testing equally. Family history of SCD was more common among CH patients [11 (6%) vs. 26 (4.2%), p=0.048]. CH patients were more likely to have a pacemaker implantation, [7 (3.8%) vs. 6 (1.0%), p=0.007] and to be treated with non-dihydropyridine calcium channel blockers [39 (21.3%) vs. 77 (12.5%), p=0.003].

No differences were found in echocardiographic or cardiac MRI parameters, or other HCM features such as syncope or abnormal blood pressure response (ABPR) at exercise according to CH (Supplementary Table 2). However, the presence of an apical aneurysm [33 (5.4%) vs. 14 (7.7%), p=0.004] and death or need for OHT were higher among HCM patients with CH [6 (3.3%) vs. 4 (0.6%), p=0.005]. MACE was also more frequent among patients with CH [12 (2.0%) vs. 9 (5.2%), p=0.026], with an unadjusted HR of 2.72 (95% CI 1.14-6.49; p=0.023), and adjusted HR of 3.46 (95% CI 1.25-9.52; p=0.016). CH patients showed a worse survival in comparison to those without CH as shown in the Kaplan-Meier curve (Log-Rank P=0.018) (Figure 2A).

**Figure 2.**
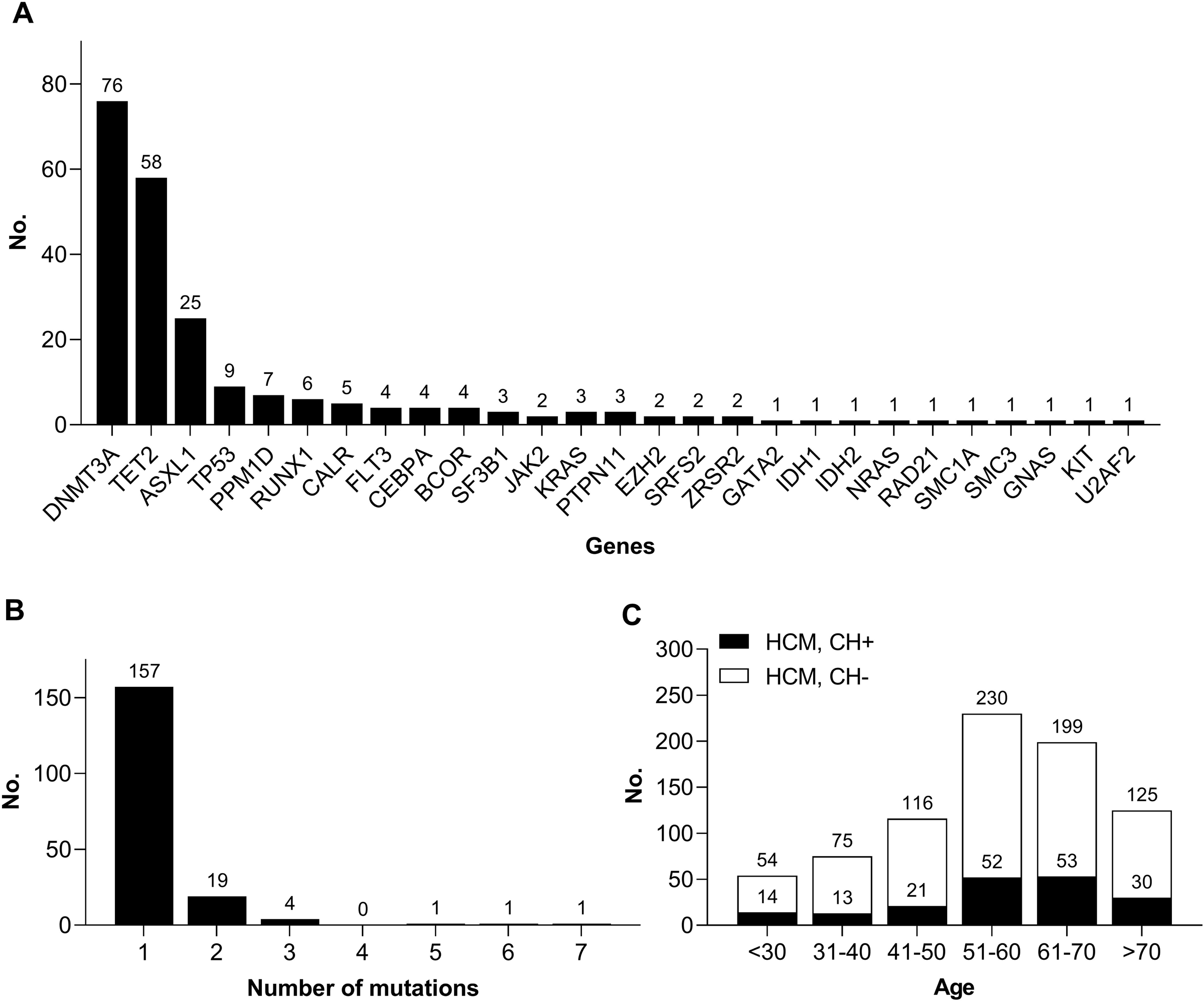
Survival in patients with HCM stratified according to CH. Panel A shows the survival according to the presence of CH among HCM patients. Panel B shows the survival according to the presence of clonal haematopoiesis in the *DNMT3A, TET2,* and *ASXL1* genes among HCM patients. Panel C shows the survival according to the presence of clonal haematopoiesis among HCM patients with germline P/LP sarcomeric variants. Panel D shows the survival according to the presence of clonal haematopoiesis in the *DNMT3A, TET2,* and *ASXL1* genes among HCM patients with germline P/LP sarcomeric variants. (CH, clonal haematopoiesis; HCM, hypertrophic cardiomyopathy; P/LP, pathogenic/likely pathogenic).

### Clonal haematopoiesis related to *DNMT3A*, *TET2* and *ASXL1*

Since *DNMT3A*, *TET2* and *ASXL1* are the most frequent and more lethal CH-associated gene mutations^9^, we performed a sub-analysis including these 3 genes and comparing to patients without CH in other genes. Patients harbouring *DNMT3A*, *TET2* and *ASXL1* mutations were older than their counterparts [58.1±14.5 vs. 55.3±14.6, p=0.046] and more likely to have a family history of SCD [9 (6.7%) vs 26 (4.2%), p=0.035]. As we note in the broader CH analysis, these patients were more likely to have hypertension, a P/LP HCM germline variant, a pacemaker and treated with a non-dihydropyridine calcium channel blocker (Supplementary Table 3).

While patients with *DNMT3A*, *TET2* and *ASXL1* CH mutations exhibited a similar HCM phenotype to those without CH (Supplementary Table 4), patients with CH showed a higher mortality or need for OHT [6 (4.4%) vs. 4 (0.6%), p<0.0001] and MACE [8 (6.2%) vs. 12 (2.0%), p=0.008]. The unadjusted HR for MACE was 3.25 (95% CI 1.32-7.98; p=0.010) and adjusted HR 3.34 (95% CI 1.33-8.42; p=0.010). Figure 2B shows the Kaplan-Meir curve with worse survival for patients with CH (Log-Rank p=0.006).

### Clonal haematopoiesis in HCM patients with germline sarcomeric gene P/LP variant

HCM patients harbouring a germline sarcomeric P/LP variant are associated with early age at diagnosis, higher MLVWT and worse outcomes.^23^ In our cohort, patients with a germline sarcomeric P/LP variant were associated with worse MACE (HR 3.1, 95% CI 1.17-8.49, p=0.022). As this subtype of HCM patients have a distinct clinical profile and are less likely to be phenocopies, we sought to investigate the CH effect in this subset of patients. Only patients with genetic screening were included [N=680 (85.1%)]. Clinical characteristics and outcomes are summarized in Supplementary Table 5 and in (Table 2 respectively. Those with CH mutations showed a trend towards a higher burden of symptoms (NYHA class II to IV) [15 (40.5%) vs. 47 (28.0%), p=0.132]. CH was also associated with a higher burden of fibrosis as reflected by LGE in >15% of the LV mass [11 (29.7%) versus 26 (15.3%), p=0.044], with an odds ratio of 2.32 (95% CI 1.00-5.38, p=0.048). A trend towards a higher degree of fibrosis in those patients with CH was also observed [15.6 % (10.4-24.6) vs. 12.2 % (7.7-17.6), p=0.068]. CH patients did show a higher frequency of ABPR at exercise [10 (62.5%) vs. 25 (29.8%), p=0.012]. Finally, CH was associated with greater mortality or need for OHT [3 (8.1%) vs. 2 (1.2%), (p=0.013)] and MACE [5 (13.5%) vs. 5 (3.0%), p=0.008], with an unadjusted HR of 5.28 (95% CI 1.51-18.4, p=0.009), and adjusted HR of 6.89 (95% CI 1.78-26.6, p=0.005). Figure 2C shows the Kaplan-Meier survival curve showing a worse survival for those with CH (log-rank p=0.003).

**Table 2.**
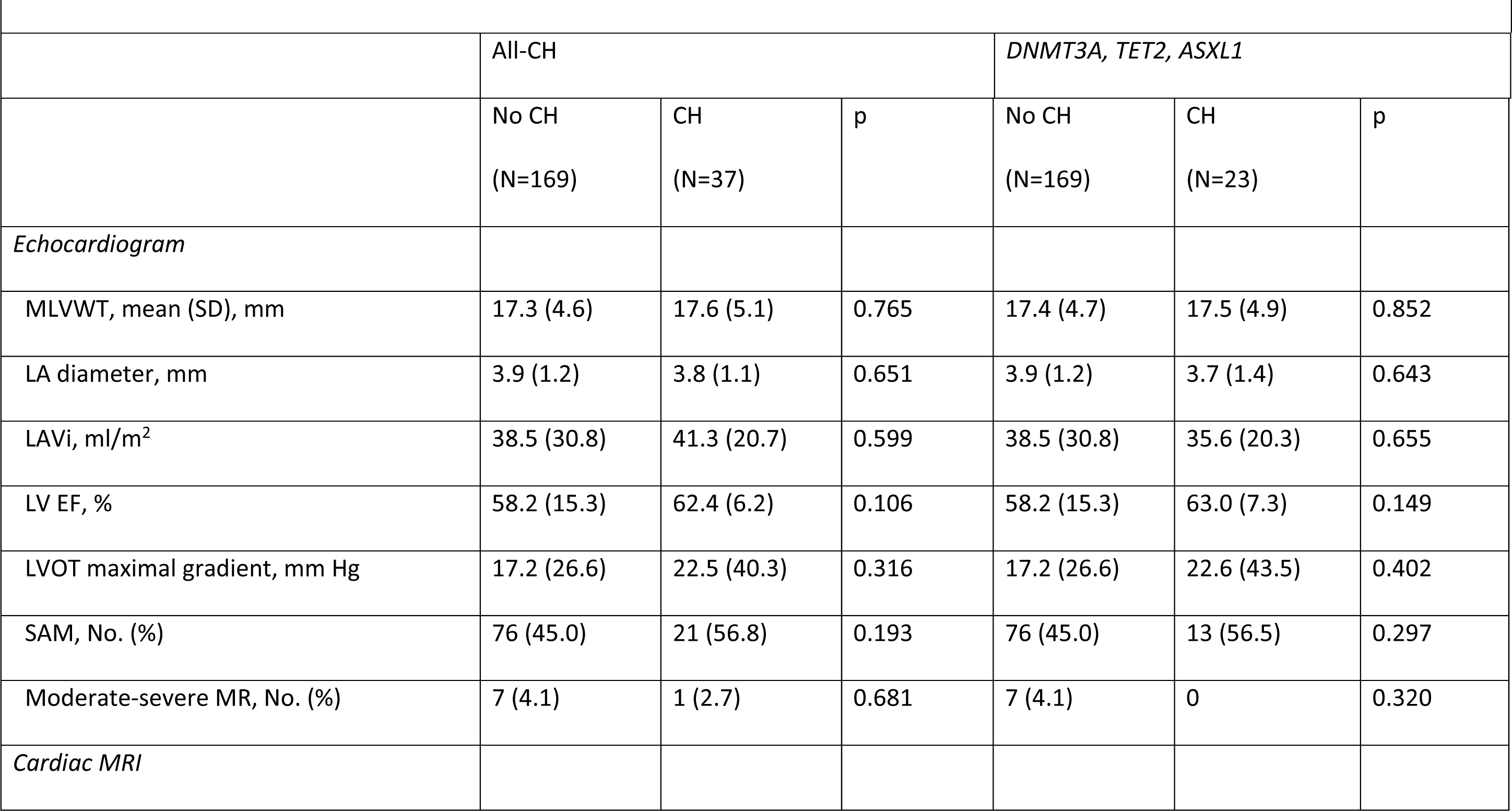
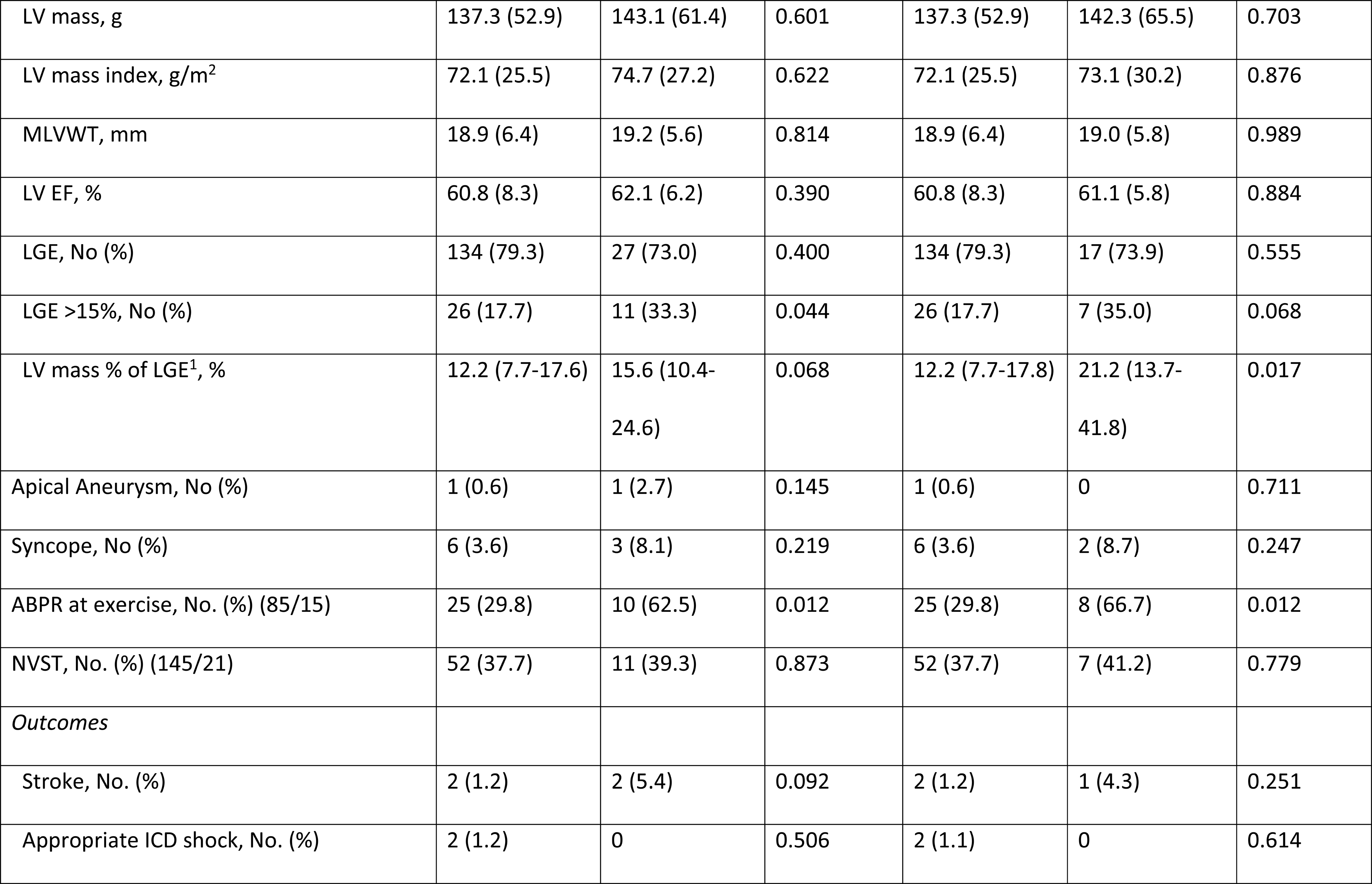
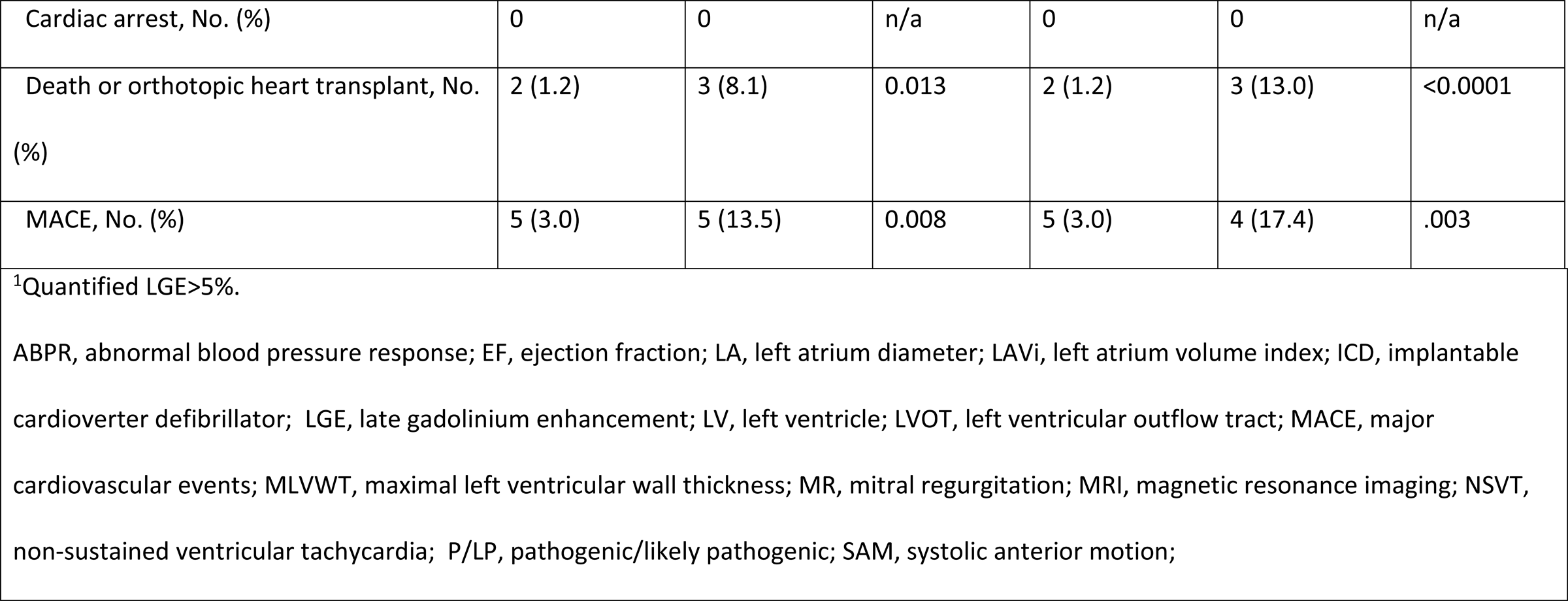
Clinical phenotype and outcomes according to CH and specific CH-related genes among HCM patients with germline P/LP sarcomeric variants.

We then evaluated the HCM phenotype and clinical outcomes in those with germline sarcomeric genes P/LP variants according to *DNMT3A*, *TET2*, and *ASXL1* CH mutations. Baseline characteristics were similar between groups (Supplementary Table 6). The amount of fibrosis assessed by LGE was higher in those with CH [21.2% (13.7-41.8) vs. 12.2% (7.7-17.6), p=0.017]. CH patients showed a higher frequency of ABPR with exercise than those without CH [8 (66.7%) vs. 25 (29.8%), p=0.012). Mortality or OHT was more common in the CH group than in those without CH [3 (13.0%) vs. 2 (1.2%), p<0.0001], as well as MACE [4 (17.4%) vs. 5 (3.0%), p=0.003]. MACE was associated with CH with an unadjusted HR of 5.28 [(95%CI 1.51-18.41) p=0.009] and for the specific *DNMT3A, TET2* and *ASXL1* CH-mutated genes had a HR of 5.76 [(95%CI 1.51-21.94), p=0.010]. Figure 2C and 2D shows the Kaplan-Meier survival curve showing a worse survival for those with CH and by specific CH mutations, respectively (log-rank p=0.007).

### Propensity score matched analysis

A propensity score matching was performed to address potential confounding factors. The group balance is shown in the Supplementary material. After the matching procedure, 514 patients without CH and 183 with CH were selected. Clinical characteristics, the HCM phenotype and outcomes are shown in Supplementary tables 7 and 8, respectively. CH was associated with MACE with an unadjusted HR of 2.78 (95% CI 1.15-6.72; p=0.023) and an adjusted HR of 2.47 (95% CI 1.01-6.02; p=0.046). We then performed a subgroup analysis according to CH and genotype. Patients with CH and germline P/LP variants were associated with higher MACE with an unadjusted HR of 11.3 (95% CI 3.28-39.4; p<0.0001) and an adjusted HR of 12.0 (95% CI 3.4-41.8; p<0.0001). Similar results were observed testing for specific CH mutations (*DNMT3A, TET2* and ASXL1) with a HR of 6.06 (95%CI 1.47-24.94; p=0.013) and with an adjusted HR of 7.31 (94% CI 1.66-32.09; p=0.008). The central figure shows the Kaplan-Meier survival curve and the forest plot for HR according to these subgroups.

### Biomarkers and cytokines/chemokines

A panel of 71 cytokines and chemokines, B-type natriuretic peptide and cardiac troponin I were evaluated in patients with a germline sarcomeric P/LP variant and levels were compared among those with (N=37) or without CH (N=169) (Figure 3). We observed significant elevated circulating levels of troponin I, IL-1ra, IL-6, IL-17F, TGF*α*, CCL21, CCL1, CCL8, and CCL17 in patients with CH. We then sought to investigate if specific CH mutations were associated with specific cytokines/chemokines. In HCM patients with germline P/LP sarcomeric variants, those with CH due to *DNMT3A* (N=16) mutations showed higher levels of IL-9 and CXCL12. *TET2* CH carriers (N=6) showed higher levels of troponin I, IL-5, IL-10, CXCL10, CXCL9, CCL4, VEFG-A, CCL21, CXCL13 and CCL1. Cytokines, chemokines, troponin I and B-type natriuretic peptide results among HCM patients with a germline P/LP sarcomeric variant and with or without CH is shown in Supplementary Table 9.

**Figure 3.**
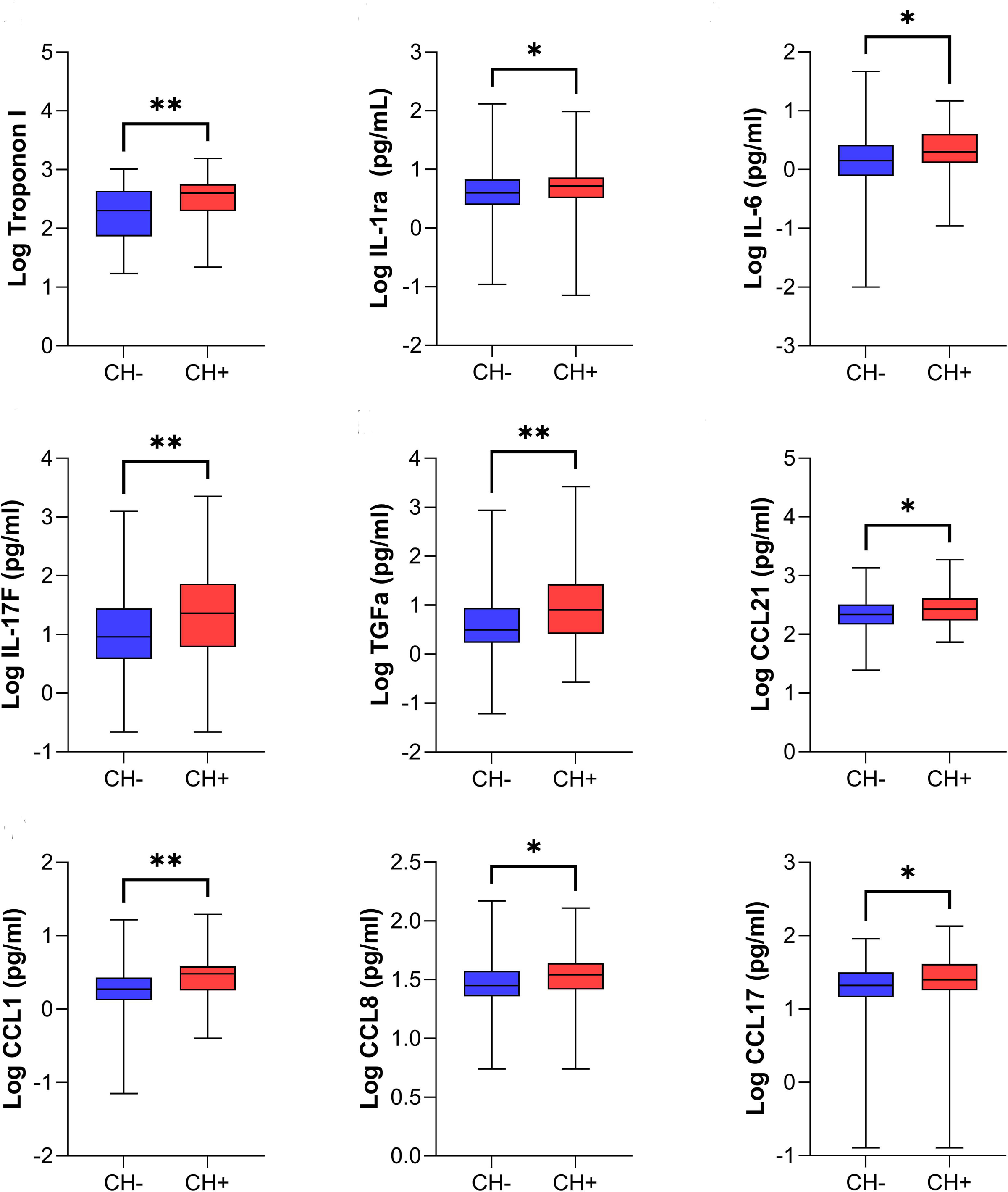
Troponin I, cytokines and chemokines levels among HCM with germline sarcomeric P/LP variant according to CH. (CH, clonal haematopoiesis; HCM, hypertrophic cardiomyopathy, P/LP, pathogenic/likely pathogenic). HCM patients with germline sarcomeric genes LP/P variants show upregulated levels of Troponin I, cytokines and chemokines.

## Discussion

The heterogeneity in HCM patients arises from mutations interacting with genetic and nongenetic factors.^24^ Our study found a higher CH prevalence in HCM patients, linked to more symptoms, ABPR during exercise, fibrosis and MACE. In addition, there were differential levels of circulating cytokines/chemokines and troponin in HCM patients with CH, highlighting a potential causal mechanism between the phenotype and outcomes. This is the first description of an association between CH prevalence, HCM phenotype and clinical outcomes.

Population studies have shown that CH prevalence ranges between 0-29% and increases with age.^11^ However, CH prevalence depends on assay sensitivity, VAF cut-off and the selected population of interest.^11,25,26^ In the present study, we found that 22.9% of HCM patients harboured CH mutations. Although most CH mutations were found in those >50 years, 25.9% of the patients <30 years also had CH in our study. Interestingly, somatic mutations in haematopoietic stem cells occurring in the first decades of life are polyclonal and have unknown long-term effects.^27,28^

Many studies have focused on the most common gene variants, such as *DNMT3A*, *TET2* and *ASXL1*^9,12,22,26,29^, present in 17% of HCM patients with CH mutations. Unsurprisingly, we found that HCM patients with these specific genes showed worse clinical phenotype and outcome, similarly to other patient cohorts.^9,12,26,29^ Experiments have shown that CH mediated by these genes were also associated with LV remodelling and interstitial fibrosis^14,16,30–32^, with evidence of macrophages migrating to the myocardium and enhancing systemic inflammation.^15,16^

HCM is characterized by asymmetric LV hypertrophy accompanied by LV outflow tract obstruction and myocardial fibrosis.^1^ While the CH impact on HCM was never explored in animal models, we observed that MLVWT was not affected by CH, even when restricting to gene-positive HCM patients. The HCM expression occurring before the acquisition of CH mutations may explain this lack of association.

Fibrosis was more common in patients with HCM who have a sarcomeric P/LP variant and *DNMT3A*, *TET2* and *ASXL1* somatic mutations. These patients also exhibited higher levels of inflammatory cytokines/chemokines, potentially contributing to increased fibrosis. This is an important finding as fibrosis is linked to SCD in patients with HCM.^33^ In addition, ABPR at exercise, a prognostic marker in HCM^34^, was also more frequent among those with CH and HCM with sarcomeric germline P/LP variants. This result potentially illustrates that CH could affect the HCM phenotype and promote adverse outcomes. Thus, these observations may reflect a ‘2-hit’ sequence of events with the first hit being the germline P/LP sarcomeric mutation followed by the second hit driven by CH.

In this study, we showed that, in subsets of HCM patients, the mortality or need for OHT is increased among CH patients even when adjusting for confounding factors, being in consonance with prior studies.^9,12,22,26,29,35^ In addition, a higher CH associated risk was observed in those with sarcomeric P/LP variants and/or specific CH genes. Our results do not address the specific CH mechanisms involved, but we hypothesize that epigenetic aging and the inflammatory milieu may drive these outcomes.

The increased inflammatory pathways recently shown in HCM^18,36^ associated with higher levels of circulating inflammatory cytokines/chemokines in CH,^9,15,16^ may have a role in driving worse outcomes. In fact, we observed higher circulating levels of several inflammatory cytokines, chemokines, and troponin I among those with CH. However, specific CH mutations may have distinct inflammatory profiles.^9,22^ We observed that *DNMT3A*-driven CH was associated with IL-9 and CXCL12. *TET2*-driven CH was associated with the higher circulating levels of a different array of cytokines/chemokines including troponin I. Our own previous work has shown that *TET2*-driven CH have been associated with a higher inflammatory milieu and a higher lethality, not seen with *DNMT3A*.^9,16,25,30^

### Limitations

There are several limitations that should be considered. Despite minimal biospecimen-to-echocardiogram timing, CH evaluation to MRI time varied (median 2.2 years, range <1 to almost 6 years). As fibrosis can progress among HCM patients^7^, it could have influenced the results. Exclusion of non-MRI patients could introduce a selection bias. The low number of events limits our conclusions especially in those without germline P/LP variants. Further cohorts needed to fully evaluate CH’s impact on HCM.

### Conclusions

The variability in the expression of the HCM phenotype could be explained by an interaction of genetic and epigenetic regulators. This study showed that the CH prevalence among HCM patients is higher than expected for a similar age. CH was associated with the presence of fibrosis and with poorer outcomes. CH is a new paradigm in HCM, impacting the phenotype and prognosis.

## Data Availability

All data produced in the present work are contained in the manuscript

## Central figure. Survival in HCM according to somatic CH-related mutations and sarcomeric germline P/LP variants

HCM patients with CH and germline sarcomeric gene LP/P variants show the worse prognosis in comparison to HCM patients without CH or without germline sarcomeric gene LP/P variants, highlighting that the presence of both risk factors warrants concern.

(CH, clonal haematopoiesis ; G, germline sarcomeric gene P/LP variants; LP, likely pathogenic; P, pathogenic).

