## Supplemental for "Clonal haematopoiesis is associated with major adverse cardiovascular events in patients with hypertrophic cardiomyopathy"

### SUPPLEMENTARY APPENDIX

**Supplementary Methods**………………………………………….……………………. Page 2

**Supplementary Statistical Analysis**……………………………………………….. Page 5

**Supplementary Table 1**. Identified Mutations in Clonal Haematopoiesis Related Genes…………………………………………………………………………….. Page 11

**Supplementary Table 2.** Clinical phenotype and outcomes according to CH among HCM patients………………………………………………………………………. Page 25

**Supplementary Table 3.** Overall characteristics of the HCM cohort and between those with or without CH in *DNMT3A, TET2* and *ASXL1*…………………………... Page 26

**Supplementary Table 4.** Clinical phenotype and outcomes in the HCM cohort and between those with or without CH in *DNMT3A, TET2* and *ASXL1*………………... Page 27

**Supplementary Table 5.** Overall characteristics of the HCM cohort with sarcomeric mutations among those with or without CH …………………………………………… Page 28

**Supplementary Table 6.** Overall characteristics of the HCM cohort with sarcomeric mutations and between those with or without CH in DNMT3A, TET2 and ASXL1…. Page 30

**Supplementary Table 7.** Overall characteristics of the HCM cohort and between those with or without CH after propensity matching…….…………………………... Page 31

**Supplementary Table 8.** Clinical phenotype and outcomes in the HCM cohort and between those with or without CH after propensity matching …….………………... Page 32

**Supplementary Table 9.** Cytokines and chemokines expression among HCM patients with sarcomeric mutations according to CH-associated gene mutations…. Page 33

#### Biospecimen analysis

The Peter Munk Cardiovascular Biobank is biorepository with associated data obtained with the consent of patients and substitute-decision makers, for potential use in future studies. Access to these resources is subject to approval from our institutional research ethics board, and studies must be reviewed by an oversight committee to ensure scientific validity. Standard operating procedures govern the processes for obtaining patient consent, specimen collection, and specimen storage. To ensure long-term preservation, aliquots of biospecimens are stored in liquid nitrogen. In the current study, buffy coat and plasma aliquots were released to Dr. Billia's laboratory after receiving institutional research ethics approval. Genomic DNA was extracted and purified from the buffy coat, and each sample was subjected to quality control to assess DNA concentration and ensure quality. Genomic DNA was then prepared according to the Ontario Institute for Cancer Research's Advanced Diagnostic Medical Laboratory's requirements for targeted sequencing. Plasma aliquots were used to perform cytokine analysis using the EVE technologies platform.

#### Clonal haematopoiesis targeted gene sequencing

We performed next-generation sequencing library construction using smMIPs to target the following 35 frequently mutated CH and myeloid malignancies genes: *ASXL1, BCOR, BRAF, CALR, CBL, CEBPA, DNMT3A, EZH2, FLT3A, GATA1, GATA2, GNAS, IDH1, IDH2, JAK2, KIT, KRAS, MPL, NRAS, PHF6, PPM1D, PTPN11, RAD21, RUNX1, SETBP1, SF3B1, SMC1A, SMC3, SRSF2, STAG2, TET2, TP53, U2AF1, WT1,* and *ZRSR2*. DNA was extracted from buffy coats, and the Illumina Novaseq platform was used to generate paired-end 150bp sequences. Each sample was sequenced twice to minimize false-positive mutation calls. We used smMIP-tools' computational pipeline to call single nucleotide variants and short insertions and deletions. This pipeline implements binomial error-rate models to calculate P-values reflective of the likelihood of true mutation calls. The average sequencing coverage across all samples was 9,950x. In all the analyses, only mutations with a variant allele frequency (VAF) of 2% were used. More broadly, the minimal number of single-strand consensus sequences for all the reported mutations was four. A 70% cut-off was required to call a consensus. Reported mutations required at least one family of size 10 reads. If a mutation was represented only by smaller families, we require those raw reads to represent at least 7 different unique molecules. Non-reference alleles sequenced in each patient sample were compared with those identical alleles in all the other unrelated technical replicates. A P-value < 0.05 after Bonferroni correction was used. To account for the potential reduction in sensitivity, only alleles with VAF < 0.05 were used to model error rates. We filtered synonymous mutations, those in non-coding regions and variants falling within interionic regions > 2bp of splicing sites. We reported non-reference alleles with Minor Allele Frequency <0.01 to mitigate the inclusion of population-level recurrent SNPs and Combined Annotation Dependent Depletion score (CADD) ≥ 10 to enrich for variants with higher predicted deleterious effects.

In addition, variants were manually inspected to mitigate the inclusion of false positives using information from smMIP-tools' output, helping to evaluate the binomial distributions' accuracy to model each allele's error rate. This information includes the number of samples detected with similar VAF and replicates that passed the model, yet their other matching replicate did not. Additional details concerning smMIP-tools output and filters can be found in the smMIP-tools' manuscript and the GitHub repository at: https://github.com/abelson-lab/smMIP-tools. The number of reads supporting the alternative allele, number of samples with VAF below the P-value threshold (0.05), and observed allele frequency compared to the second lowest allele frequency.

**Cytokine analysis**

The Luminex™ 200 system, provided by Eve Technologies Corp. in Alberta, Canada, was used to analyse plasma samples. The Human Cytokine 48-Plex Discovery Assay® (MilliporeSigma, Massachusetts, USA) was employed in accordance with the manufacturer's instructions. The 71-plex was comprised of various cytokines and chemokines, including CCL21, sCD40L, EGF, CCL-11, FGF-2, FLT3, CX3CL1, G-CSF, GM-CSF, CXCL1, IFNα2, IFNγ, IL-1α, IL-1β, IL-1RA, IL-2, IL-3, IL-4, IL-5, IL-6, IL-7, IL-8, IL-9, IL-10, IL-12p40, IL-12p70, IL-13, IL-15, IL-17A, IL-17E/IL-25, IL-17F, IL-18, IL-22, IL-27, CXCL10, CCL2, CCL7, CSCF1, CXCL12, CXCL9, CCL3, CCL4, PDGF-AA, PDGF-AA/BB, CCL5, TGF-α, TNF-α, TNF-β, VEGF-A, CXCL13, CCL27, CXCL5, CCL24, CCL26, CCL1, IL-16, IL-20, IL-21, IL-23, IL-28[a], IL-33, LIF, CCL8, CCL13, CCL15, SCF, CXCL12, CCL17, TPO, TRAIL, and TSLP. In addition to the aforementioned cytokines/chemokines, levels of brain natriuretic peptide (BNP) and cardiac troponin I (cTnI) were measured, as these biomarkers play a key role in the inflammatory response.

**Statistical Analysis**

Continuous data were evaluated for normality with Shapiro-Wilk test and histogram analysis and presented as mean ± standard deviation or median (interquartile range) accordingly. Categorical variables were presented as total count (relative frequency). A Student's T test was used to compare continuous variables with normal distribution or with Mann-Whitney test for those with non-parametric distribution. ANOVA test was used to compare continuous variables among >2 groups. Chi-square test or Fisher's exact test were used for comparing categorical variables between groups accordingly. Fibrosis analysis with quantitative measurement included only patients with available images for quantification, all others were excluded. Because the limitations for quantifying extreme low amount of fibrosis, the quantitative LGE fibrosis analysis was performed only in patients with at least 5% of LGE on the LV. Survival among those with CH or without CH were evaluated by a Kaplan-Meier method and survival differences evaluated by the log-rank test. We used Cox proportional hazards regressions to compare survival rates between different groups. We included age as a covariate in the adjusted Cox proportional hazards regressions due to its significant interaction with CH. We also tested the proportional hazard assumptions through visual inspection and Schoenfeld residuals. To control possible confounders between patients with or without CH, we did a propensity score matching with up to 1:3 ratio using a nearest neighbor method including the following variables in the matching: sex, family history of SCD, hypertension, diabetes, MLVWT, LVOT maximum gradient, NYHA functional class, and NSVT. The balance between groups was assessed by standardized differences. As a sensitivity analysis, we also tested those with CH only in the three most common CH genes (*DNMT3A, TET2* and *ASXL1*). We used a statistical significance of 0.05 for all analyses and a two-sided p-value. All analyses were performed using SPSS, version 25.0 (SPSS Inc., NY, USA).

**Propensity Matching Analysis**

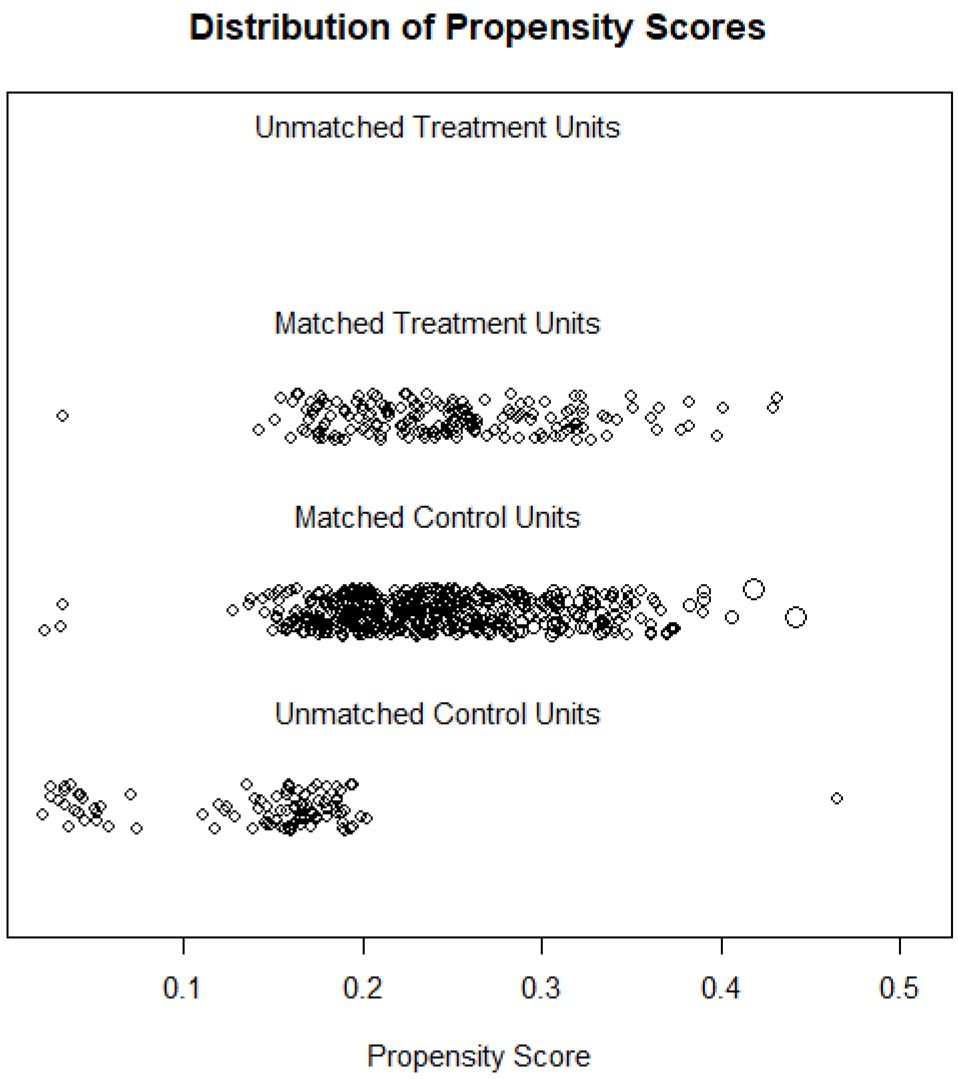

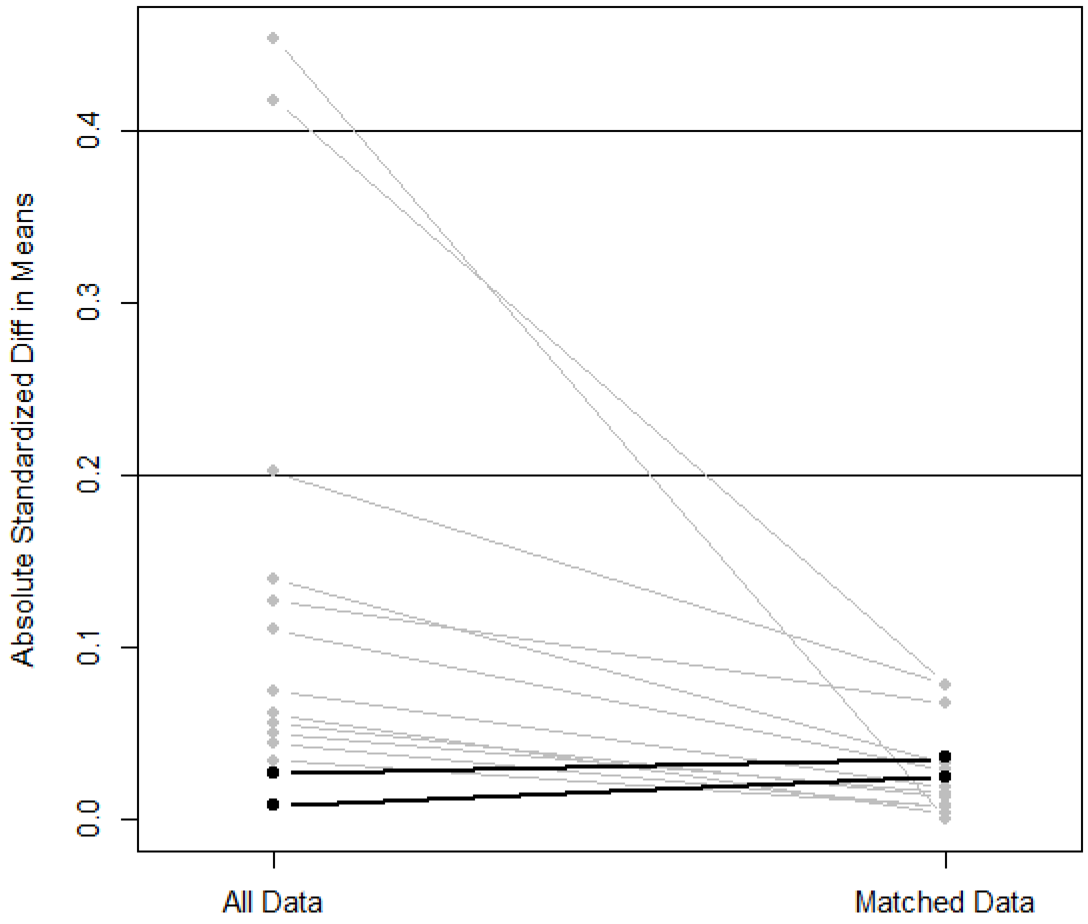

Absolute Standardized Diff in Means
0.0
0.1
0.2
0.3
0.4
All Data
Matched Data

100

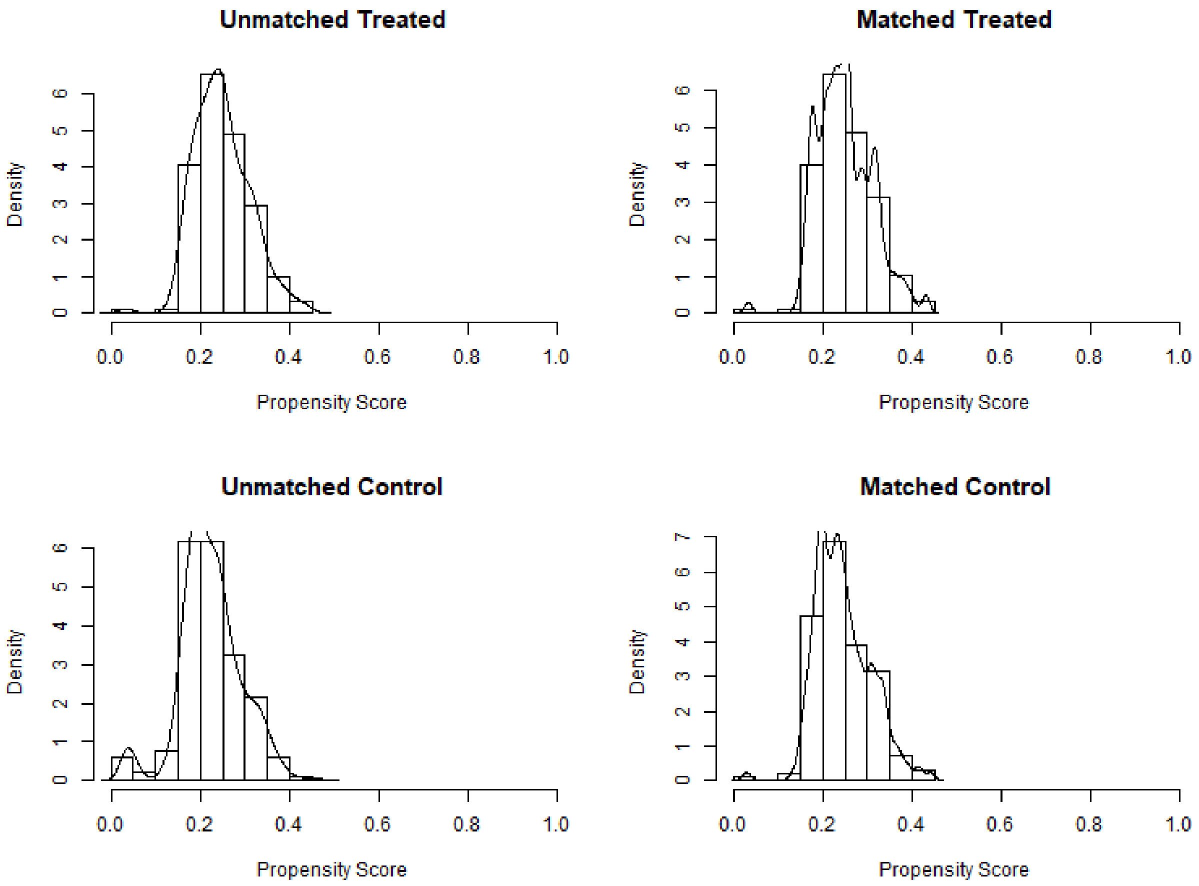
Unmatched Treated
Matched Treated
Unmatched Control
Matched Control
Density
Propensity Score
0.0
0.2
0.4
0.6
0.8
1.0
0
1
2
3
4
5
6
7

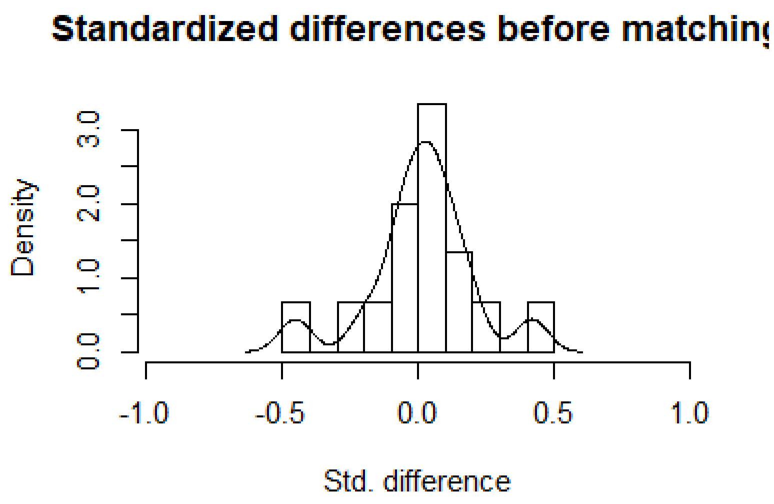
Standardized differences before matchin
Density
0.0
1.0
2.0
3.0
-1.0
-0.5
0.0
0.5
1.0
Std. difference

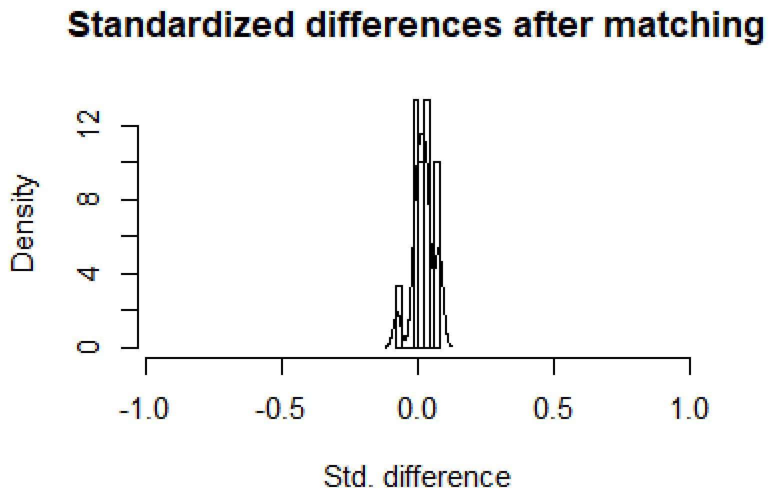
Standardized differences after matching
Density
0
4
8
12
-1.0
-0.5
0.0
0.5
1.0
Std. difference

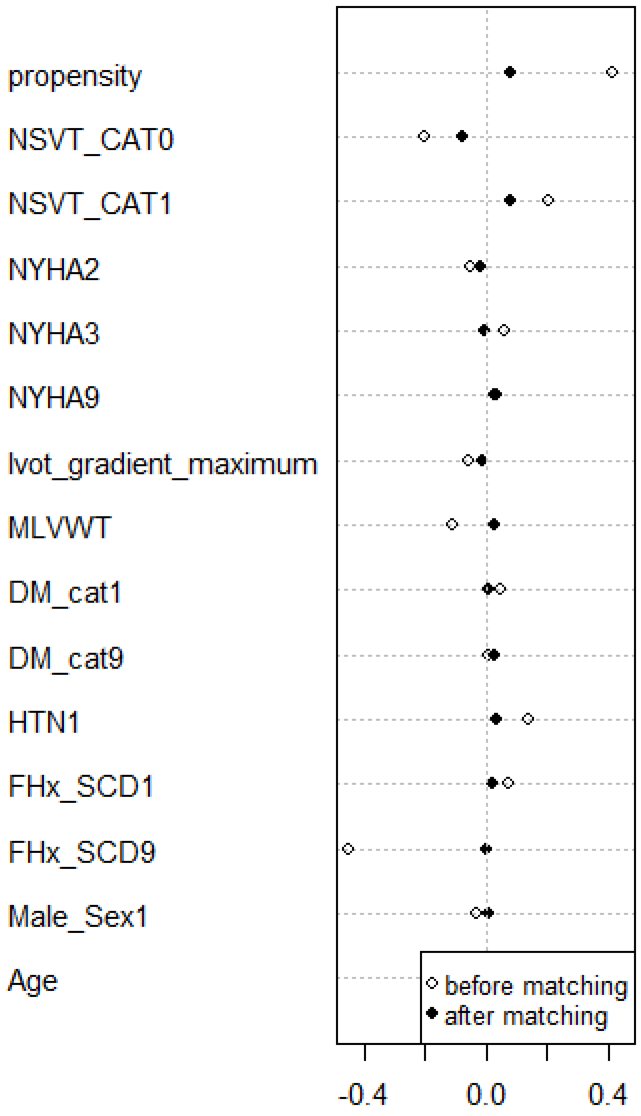
propensity
NSVT_CAT0
NSVT_CAT1
NYHA2
NYHA3
NYHA9
lvot_gradient_maximum
MLVWT
DM_cat1
DM_cat9
HTN1
FHx_SCD1
FHx_SCD9
Male_Sex1
Age
before matching
after matching
-0.4
0.0
0.4

**Supplementary Table 1.** Identified Mutations in Clonal Haematopoiesis Related Genes.

| Sample ID | **Gene** | **Chromosome** | **Start position** | **End position** | **Variant type** | **Variant type** | **Ref. Allele** | **Alt. Allele** | **Protein change** | **Alt. reads** | **Ref. reads** | **VAF** |
|---|---|---|---|---|---|---|---|---|---|---|---|---|
| HCM0005 | DNMT3A | chr2 | 25468120 | 25468120 | splice_donor_variant | SNV | A | G | c.1554+2T>C | 178 | 13892 | 0.0128 |
| HCM0005 | DNMT3A | chr2 | 25464533 | 25464535 | Frame_Shift_Del | SNV | GTA | - | Tyr660fs | 307 | 10379 | 0.0296 |
| HCM0005 | TET2 | chr4 | 106182964 | 106182964 | missense_variant | SNV | C | T | Pro1335Ser | 20229 | 42368 | 0.4775 |
| HCM0007 | EZH2 | chr7 | 148516734 | 148516734 | missense_variant | SNV | G | T | Thr318Lys | 420 | 23341 | 0.0180 |
| HCM0008 | EZH2 | chr7 | 148506477 | 148506477 | missense_variant | SNV | C | A | Val679Leu | 152 | 8234 | 0.0185 |
| HCM0011 | SF3B1 | chr2 | 198267359 | 198267359 | missense_variant | SNV | C | A | Lys666Asn | 400 | 14653 | 0.0273 |
| HCM0011 | TET2 | chr4 | 106156478 | 106156478 | missense_variant | SNV | C | T | Ser460Phe | 4859 | 9549 | 0.5088 |
| HCM0011 | SRSF2 | chr17 | 74732959 | 74732959 | missense_variant | SNV | G | A | Pro95Leu | 344 | 4816 | 0.0714 |
| HCM0012 | TET2 | chr4 | 106156060 | 106156060 | Frame_Shift_Del | Indel | C | - | Gln321fs | 7705 | 18861 | 0.4085 |
| HCM0014 | ASXL1 | chr20 | 31021127 | 31021127 | missense_variant | SNV | G | T | Gly376Cys | 303 | 19057 | 0.0159 |
| HCM0017 | DNMT3A | chr2 | 25457242 | 25457242 | missense_variant | SNV | C | T | Arg882His | 3827 | 22398 | 0.1709 |
| HCM0017 | DNMT3A | chr2 | 25463223 | 25463223 | missense_variant | SNV | T | A | Asn757Ile | 1000 | 16492 | 0.0606 |
| HCM0018 | TET2 | chr4 | 106196291 | 106196291 | stop_gained | SNV | C | T | Gln1542* | 267 | 36736 | 0.0073 |
| HCM0020 | RUNX1 | chr21 | 36253004 | 36253004 | missense_variant | SNV | C | A | Ala120Ser | 152 | 27452 | 0.0055 |
| HCM0020 | TET2 | chr4 | 106196604 | 106196604 | Frame_shift_indel | Indel | T | - | Leu1646fs | 5139 | 38744 | 0.1326 |
| HCM0021 | EZH2 | chr7 | 148512061 | 148512061 | stop_gained | SNV | G | T | Cys539* | 552 | 17620 | 0.0313 |
| HCM0022 | TET2 | chr4 | 106180896 | 106180896 | missense_variant | SNV | G | T | p.Lys1308Asn | 452 | 21630 | 0.0209 |
| HCM0023 | TET2 | chr4 | 106158419 | 106158419 | Frame_shift_indel | Indel | C | - | Ser1107fs | 458 | 20575 | 0.0223 |
| HCM0024 | DNMT3A | chr2 | 25457168 | 25457168 | stop_gained | SNV | C | A | Glu907* | 314 | 10028 | 0.0313 |
| HCM0025 | CALR | chr19 | 13054627 | 13054627 | Frame_shift_ins | Indel | A | + | Glu386fs | 14214 | 35697 | 0.3982 |
| HCM0025 | TET2 | chr4 | 106155921 | 106155921 | Frame_shift_indel | Indel | C | - | Asn275fs | 359 | 21434 | 0.0167 |
| HCM0025 | DNMT3A | chr2 | 25466797 | 25466797 | missense_variant | SNV | C | A | Val636Leu | 92 | 6408 | 0.0144 |
| HCM0025 | DNMT3A | chr2 | 25464552 | 25464552 | missense_variant | SNV | C | T | Gly654Asp | 393 | 14548 | 0.0270 |
| HCM0025 | TET2 | chr4 | 106180895 | 106180895 | missense_variant | SNV | A | C | Lys1308Thr | 378 | 29716 | 0.0127 |
| HCM0027 | SF3B1 | chr2 | 198267371 | 198267371 | missense_variant | SNV | G | C | His662Gln | 312 | 13432 | 0.0232 |
| HCM0030 | PPM1D | chr17 | 58740541 | 58740541 | Frame_shift_indel | Indel | G | - | Thr483fs | 406 | 18258 | 0.0222 |

| | | | | | | | | | | | | |
|---|---|---|---|---|---|---|---|---|---|---|---|---|
| HCM0031 | KIT | chr4 | 55599308 | 55599308 | missense_variant | SNV | G | T | Gly812Cys | 232 | 23427 | 0.0099 |
| HCM0031 | WT1 | chr11 | 32417827 | 32417827 | missense_variant | SNV | G | T | Gln414Lys | 194 | 14291 | 0.0136 |
| HCM0033 | TET2 | chr4 | 106157205 | 106157205 | missense_variant | SNV | C | A | His702Gln | 431 | 16604 | 0.0260 |
| HCM0034 | DNMT3A | chr2 | 25469028 | 25469028 | splice_donor_variant | SNV | C | A | c.1429+1G>T | 258 | 7800 | 0.0331 |
| HCM0034 | ASXL1 | chr20 | 31022442 | 31022442 | Frame_shift_indel | Indel | G | + | Gly646fs | 220 | 24746 | 0.0089 |
| HCM0034 | DNMT3A | chr2 | 25468129 | 25468129 | Frame_shift_indel | Indel | T | - | Asn516fs | 500 | 47981 | 0.0104 |
| HCM0040 | DNMT3A | chr2 | 25469029 | 25469029 | missense_variant | SNV | C | T | Glu477Lys | 238 | 8332 | 0.0286 |
| HCM0041 | DNMT3A | chr2 | 25464433 | 25464433 | missense_variant | SNV | G | A | His694Tyr | 423 | 15969 | 0.0265 |
| HCM0045 | SF3B1 | chr2 | 198267360 | 198267360 | missense_variant | SNV | T | A | Lys666Met | 6055 | 23213 | 0.2608 |
| HCM0046 | DNMT3A | chr2 | 25463236 | 25463238 | Frame_shift_indel | Indel | AGA | - | Trp753fs | 760 | 60860 | 0.0125 |
| HCM0046 | SMC1A | chrX | 53432728 | 53432728 | missense_variant | SNV | G | T | Thr569Asn | 214 | 21179 | 0.0101 |
| HCM0047 | DNMT3A | chr2 | 25457192 | 5457192 | Frame_shift_indel | SNV | G | - | Arg899fs | 345 | 51495 | 0.0067 |
| HCM0048 | TET2 | chr4 | 106180904 | 106180904 | missense_variant | SNV | T | A | Leu1311Gln | 255 | 14309 | 0.0178 |
| HCM0048 | TET2 | chr4 | 106182916 | 106182916 | stop_gained | SNV | G | T | c.3955G>T | 352 | 12467 | 0.0282 |
| HCM0050 | DNMT3A | chr2 | 25468121 | 25468121 | splice_donor_variant | SNV | C | T | c.1554+1G>A | 217 | 14985 | 0.0145 |
| HCM0051 | DNMT3A | chr2 | 25467503 | 25467504 | Frema_shift_indel | Indel | CA | - | Ala525fs | 1069 | 65822 | 0.0162 |
| HCM0052 | DNMT3A | chr2 | 25463170 | 25463170 | splice_donor_variant | SNV | C | T | c.2322+1G> | 2773 | 15635 | 0.1774 |
| HCM0052 | TET2 | chr4 | 106164073 | 106164073 | missense_variant | SNV | A | G | Ile1195Val | 2163 | 4324 | 0.5002 |
| HCM0054 | DNMT3A | chr2 | 25463568 | 25463568 | missense_variant | SNV | A | G | Ile705Thr | 712 | 11656 | 0.0611 |
| HCM0054 | PTPN11 | chr12 | 112926261 | 112926261 | missense_variant | SNV | G | A | Arg465Gln | 258 | 14578 | 0.0177 |
| HCM0055 | ASXL1 | chr20 | 31022441 | 31022441 | Frame_shift_ins | Indel | A | + | Gly643fs | 381 | 21189 | 0.0180 |
| HCM0055 | DNMT3A | chr2 | 25463289 | 25463289 | missense_variant | SNV | T | C | Tyr735Cys | 490 | 16573 | 0.0296 |
| HCM0056 | TET2 | chr4 | 106190861 | 106190861 | Frame_shift_indel | Indel | A | + | His1380fs | 566 | 15921 | 0.0356 |
| HCM0059 | TET2 | chr4 | 106164778 | 106164778 | stop_gained | SNV | C | T | Arg1216* | 299 | 14562 | 0.0205 |
| HCM0060 | ASXL1 | chr20 | 31022770 | 31022770 | missense_variant | SNV | C | A | Ala752Asp | 459 | 24540 | 0.0162 |
| HCM0062 | EZH2 | chr7 | 148504787 | 148504787 | missense_variant | SNV | G | C | Ala736Gly | 312 | 18322 | 0.0170 |
| HCM0068 | ASXL1 | chr20 | 31022851 | 31022851 | missense_variant | SNV | C | T | Pro779Leu | 20270 | 46934 | 0.4319 |
| HCM0072 | ASXL1 | chr20 | 31021568 | 31021568 | stop_gained | SNV | A | T | Lys523* | 4233 | 38559 | 0.1098 |
| HCM0074 | CBL | chr11 | 119148927 | 119148928 | Frame_shift_indel | indel | AT | - | Ile383fs | 328 | 72978 | 0.0045 |
| HCM0074 | CBL | chr11 | 119148967 | 119148967 | missense_variant | SNV | G | A | Cys396Tyr | 394 | 116397 | 0.0034 |
| HCM0076 | DNMT3A | chr2 | 25462068 | 25462068 | missense_variant | SNV | A | G | Ile780Thr | 719 | 9119 | 0.0788 |

| | | | | | | | | | | | | |
|---|---|---|---|---|---|---|---|---|---|---|---|---|
| HCM0076 | TET2 | chr4 | 106157848 | 106157848 | stop_gained | SNV | C | T | Gln917* | 161 | 14437 | 0.0112 |
| HCM0076 | DNMT3A | chr2 | 25467498 | 25467498 | stop_gained | SNV | G | C | Tyr526* | 4724 | 39043 | 0.1210 |
| HCM0076 | TET2 | chr4 | 106196303 | 106196303 | stop_gained | SNV | C | T | Gln1546* | 152 | 19900 | 0.0076 |
| HCM0079 | DNMT3A | chr2 | 25467449 | 25467449 | missense_variant | SNV | C | T | Gly543Ser | 412 | 6871 | 0.0600 |
| HCM0081 | DNMT3A | chr2 | 25464573 | 25464573 | missense_variant | SNV | A | G | Leu647Pro | 191 | 9178 | 0.0208 |
| HCM0084 | ASXL1 | chr20 | 31021514 | 31021514 | missense_variant | SNV | G | T | Asp505Tyr | 604 | 59195 | 0.0102 |
| HCM0085 | DNMT3A | chr2 | 25468154 | 25468154 | missense_variant | SNV | G | T | Leu508Ile | 358 | 13046 | 0.0274 |
| HCM0086 | ASXL1 | chr20 | 31022441 | 31022441 | Frame_shift_ins | Indel | A | + | Gly643fs | 2127 | 29087 | 0.0731 |
| HCM0086 | TET2 | chr4 | 106194022 | 106194022 | Frame_shift_indel | Indel | C | - | Pro1496fs | 453 | 38039 | 0.0119 |
| HCM0086 | EZH2 | chr7 | 148506181 | 148506181 | missense_variant | SNV | T | G | Glu726Ala | 177 | 26475 | 0.0067 |
| HCM0086 | TET2 | chr4 | 106197285 | 106197285 | missense_variant | SNV | T | C | Ile1873Thr | 370 | 15979 | 0.0232 |
| HCM0088 | SMC3 | chr10 | 112350199 | 112350199 | missense_variant | SNV | C | A | Asn513Lys | 113 | 21782 | 0.0052 |
| HCM0088 | DNMT3A | chr2 | 25464544 | 25464544 | missense_variant | SNV | C | T | Val657Met | 192 | 9627 | 0.0199 |
| HCM0089 | DNMT3A | chr2 | 25466812 | 25466812 | missense_variant | SNV | T | A | Arg631Trp | 77 | 2316 | 0.0332 |
| HCM0090 | DNMT3A | chr2 | 25470028 | 25470028 | splice_acceptor_variant | SNV | C | T | c.1015-1G>A | 413 | 2460 | 0.1679 |
| HCM0091 | DNMT3A | chr2 | 25457242 | 25457242 | missense_variant | SNV | C | T | Arg882His | 1630 | 62332 | 0.0262 |
| HCM0092 | WT1 | chr11 | 32413611 | 32413611 | splice_acceptor_variant | SNV | C | A | c.1355-1G>A | 286 | 26902 | 0.0106 |
| HCM0092 | TP53 | chr17 | 7578457 | 7578457 | missense_variant | SNV | C | T | Arg158His | 353 | 9399 | 0.0376 |
| HCM0093 | DNMT3A | chr2 | 25467134 | 25467134 | Frame_shift_indel | Indel | A | - | Trp581fs | 422 | 12006 | 0.0351 |
| HCM0098 | DNMT3A | chr2 | 25462068 | 25462068 | missense_variant | SNV | A | G | Ile780Thr | 449 | 11907 | 0.0377 |
| HCM0108 | DNMT3A | chr2 | 25463308 | 25463308 | missense_variant | SNV | G | A | Arg729Trp | 1524 | 11737 | 0.1298 |
| HCM0109 | TET2 | chr4 | 106155920 | 106155920 | Frame_shift_indel | Indel | T | - | Ile274fs | 576 | 17986 | 0.0320 |
| HCM0112 | CBL | chr11 | 119148991 | 119148991 | missense_variant | SNV | G | A | Cys404Tyr | 952 | 110413 | 0.0086 |
| HCM0113 | EZH2 | chr7 | 148511205 | 148511205 | missense_variant | SNV | C | A | Arg566Leu | 273 | 15856 | 0.0172 |
| HCM0113 | SMC1A | chrX | 53432580 | 53432580 | missense_variant | SNV | G | A | Arg586Trp | 252 | 13519 | 0.0186 |
| HCM0117 | BCOR | chrX | 39932878 | 39932878 | missense_variant | SNV | G | A | Pro574Leu | 587 | 17802 | 0.0330 |
| HCM0117 | SMC1A | chrX | 53432316 | 53432316 | missense_variant | SNV | G | T | Ala640Glu | 600 | 32521 | 0.0184 |
| HCM0120 | EZH2 | chr7 | 148511146 | 148511146 | missense_variant | SNV | C | A | Asp586Tyr | 384 | 19732 | 0.0195 |
| HCM0120 | DNMT3A | chr2 | 25463197 | 25463197 | Frame_shift_indel | Indel | T | + | Asp765fs | 513 | 31817 | 0.0161 |
| HCM0128 | DNMT3A | chr2 | 25462023 | 25462023 | stop_gained | SNV | C | T | Trp795* | 440 | 12576 | 0.0350 |
| HCM0128 | TP53 | chr17 | 7577577 | 7577577 | missense_variant | SNV | T | C | Asn235Ser | 14413 | 32056 | 0.4496 |

| | | | | | | | | | | | | |
|---|---|---|---|---|---|---|---|---|---|---|---|---|
| HCM0129 | DNMT3A | chr2 | 25467448 | 25467448 | missense_variant | SNV | C | G | Gly543Ala | 294 | 21022 | 0.0140 |
| HCM0130 | PTPN11 | chr12 | 112910842 | 112910842 | missense_variant | SNV | C | A | Pro284His | 429 | 8785 | 0.0488 |
| HCM0131 | DNMT3A | chr2 | 25463550 | 25463550 | missense_variant | SNV | T | A | Asn711Ile | 283 | 15135 | 0.0187 |
| HCM0134 | DNMT3A | chr2 | 25457185 | 25457185 | missense_variant | SNV | A | G | Leu901Pro | 1809 | 30520 | 0.0593 |
| HCM0136 | PTPN11 | chr12 | 112926270 | 112926270 | missense_variant | SNV | C | A | Thr468Lys | 204 | 37862 | 0.0054 |
| HCM0137 | TP53 | chr17 | 7577535 | 7577535 | missense_variant | SNV | C | G | Arg249Thr | 309 | 9490 | 0.0326 |
| HCM0142 | RAD21 | chr8 | 117866675 | 117866675 | missense_variant | SNV | G | T | Leu324Ile | 376 | 29610 | 0.0127 |
| HCM0142 | DNMT3A | chr2 | 25467523 | 25467523 | splice_acceptor_variant | SNV | T | C | c.1555-2A>G | 617 | 9133 | 0.0676 |
| HCM0144 | DNMT3A | chr2 | 25467160 | 25467168 | Frame_shift_Indel | Indel | GCAGCCCCC | - | Ala571fs | 260 | 5857 | 0.0444 |
| HCM0146 | ZRSR2 | chrX | 15836710 | 15836710 | missense_variant | SNV | G | T | Val258Phe | 355 | 15383 | 0.0231 |
| HCM0148 | TET2 | chr4 | 106157977 | 106157977 | stop_gained | SNV | C | T | Gln960* | 230 | 21673 | 0.0106 |
| HCM0148 | PPM1D | chr17 | 58740726 | 58740726 | Frame_shift_Indel | Indel | G | + | Leu546fs | 110 | 28197 | 0.0039 |
| HCM0152 | SMC1A | chrX | 53432780 | 53432780 | missense_variant | SNV | T | A | Thr552Ser | 155 | 26176 | 0.0059 |
| HCM0153 | DNMT3A | chr2 | 25459876 | 25459876 | splice_acceptor_variant | SNV | T | C | c.2409-2A>G | 436 | 23854 | 0.0183 |
| HCM0154 | ASXL1 | chr20 | 31021610 | 31021610 | missense_variant | SNV | G | A | Glu537Lys | 3275 | 9640 | 0.3397 |
| HCM0157 | JAK2 | chr9 | 5073770 | 5073770 | missense_variant | SNV | G | T | Val617Phe | 36458 | 42724 | 0.8533 |
| HCM0161 | TET2 | chr4 | 106164922 | 106164922 | missense_variant | SNV | G | A | Ala1264Thr | 16646 | 32453 | 0.5129 |
| HCM0162 | DNMT3A | chr2 | 25463302 | 25463302 | missense_variant | SNV | A | G | Phe731Leu | 188 | 18792 | 0.0100 |
| HCM0162 | TET2 | chr4 | 106155749 | 106155749 | Frame_shift_Indel | Indel | C | - | Val218fs | 88 | 23738 | 0.0037 |
| HCM0162 | DNMT3A | chr2 | 25467484 | 25467484 | missense_variant | SNV | T | C | Asp531Gly | 83 | 24137 | 0.0034 |
| HCM0164 | DNMT3A | chr2 | 25459803 | 25459803 | splice_donor_variant | SNV | A | C | c.2478+2T>G | 135 | 21639 | 0.0062 |
| HCM0167 | DNMT3A | chr2 | 25467114 | 25467114 | Frame_shift_Indel | Indel | C | - | His588fs | 208 | 14221 | 0.0146 |
| HCM0172 | CEBPA | chr19 | 33792343 | 33792343 | Frame_shift_Indel | Indel | C | - | Lys326fs | 140 | 5728 | 0.0244 |
| HCM0177 | DNMT3A | chr2 | 25464454 | 25464454 | missense_variant | SNV | C | T | Val687Ile | 206 | 7834 | 0.0263 |
| HCM0179 | CBL | chr11 | 119149241 | 119149241 | missense_variant | SNV | C | T | Pro417Ser | 87 | 13561 | 0.0064 |
| HCM0180 | ASXL1 | chr20 | 31022925 | 31022925 | stop_gained | SNV | G | T | Gly804* | 898 | 55254 | 0.0163 |
| HCM0182 | TET2 | chr4 | 106196483 | 106196483 | missense_variant | SNV | G | C | Gly1606Arg | 8244 | 19054 | 0.4327 |
| HCM0184 | ASXL1 | chr20 | 31024736 | 31024736 | missense_variant | SNV | C | A | Asp1407Glu | 204 | 23199 | 0.0088 |
| HCM0185 | ASXL1 | chr20 | 31023083 | 31023083 | stop_gained | SNV | C | A | Cys856* | 901 | 14345 | 0.0628 |
| HCM0185 | WT1 | chr11 | 32414227 | 32414227 | stop_gained | SNV | G | A | Gln447* | 68 | 15317 | 0.0044 |
| HCM0191 | PHF6 | chrX | 133547976 | 133547976 | missense_variant | SNV | G | T | Ala237Ser | 255 | 19063 | 0.0134 |

| | | | | | | | | | | | | |
|---|---|---|---|---|---|---|---|---|---|---|---|---|
| HCM0192 | DNMT3A | chr2 | 25468154 | 25468154 | Frame_shift_Indel | Indel | G | - | Ala571fs | 1560 | 11024 | 0.1415 |
| HCM0196 | ASXL1 | chr20 | 31022443 | 31022443 | missense_variant | SNV | G | C | Gly643Ala | 20049 | 35592 | 0.5633 |
| HCM0202 | DNMT3A | chr2 | 25464540 | 25464540 | missense_variant | SNV | T | A | Asp658Val | 2203 | 19119 | 0.1152 |
| HCM0204 | DNMT3A | chr2 | 25463297 | 25463301 | Frame_shift_Indel | Indel | AAAGA | - | Phe731fs | 1965 | 9531 | 0.2062 |
| HCM0209 | ASXL1 | chr20 | 31022443 | 31022443 | missense_variant | SNV | G | T | Gly643Val | 38583 | 61794 | 0.6244 |
| HCM0212 | PPM1D | chr17 | 58740706 | 58740706 | Frame_shift_Indel | Indel | A | - | Leu538fs | 330 | 14124 | 0.0234 |
| HCM0212 | TET2 | chr4 | 106157737 | 106157737 | missense_variant | SNV | C | A | His880Asn | 294 | 16706 | 0.0176 |
| HCM0216 | ASXL1 | chr20 | 31021511 | 31021511 | missense_variant | SNV | C | A | Pro504Thr | 600 | 28012 | 0.0214 |
| HCM0218 | TP53 | chr17 | 7578406 | 7578406 | missense_variant | SNV | C | T | Arg175His | 471 | 10794 | 0.0436 |
| HCM0219 | ZRSR2 | chrX | 15833879 | 15833879 | stop_gained | SNV | C | T | Gln213* | 54 | 15766 | 0.0034 |
| HCM0220 | DNMT3A | chr2 | 25462023 | 25462023 | stop_gained | SNV | C | T | Trp795* | 166 | 16704 | 0.0099 |
| HCM0221 | STAG2 | chrX | 123184091 | 123184091 | Frame_shift_Indel | Indel | A | - | Lys317fs | 106 | 33226 | 0.0032 |
| HCM0221 | DNMT3A | chr2 | 25463182 | 25463182 | stop_gained | SNV | G | A | Arg771* | 818 | 28317 | 0.0289 |
| HCM0223 | DNMT3A | chr2 | 25463508 | 25463508 | splice_donor_variant | SNV | C | T | c.2173+1G>A | 278 | 21358 | 0.0130 |
| HCM0223 | DNMT3A | chr2 | 25467492 | 25467492 | stop_gained | SNV | G | C | Tyr528* | 279 | 27602 | 0.0101 |
| HCM0224 | TET2 | chr4 | 106155916 | 106155916 | stop_gained | SNV | C | T | Gln273* | 180 | 15242 | 0.0118 |
| HCM0226 | TET2 | chr4 | 106156436 | 106156436 | stop_gained | SNV | T | G | Leu446* | 461 | 12659 | 0.0364 |
| HCM0229 | DNMT3A | chr2 | 25457242 | 25457242 | missense_variant | SNV | C | T | Arg882His | 2552 | 46345 | 0.0551 |
| HCM0229 | TET2 | chr4 | 106164086 | 106164086 | splice_donor_variant | SNV | T | A | c.3594+2T>A | 298 | 15076 | 0.0198 |
| HCM0229 | TET2 | chr4 | 106196662 | 106196662 | missense_variant | SNV | C | A | Asp1665Glu | 12111 | 29536 | 0.4100 |
| HCM0230 | KRAS | chr12 | 25380347 | 25380347 | splice_acceptor_variant | SNV | C | A | c.112-1G>T | 262 | 12166 | 0.0215 |
| HCM0232 | PPM1D | chr17 | 58740471 | 58740471 | Frame_shift_Indel | Indel | A | - | Glu459fs | 282 | 26811 | 0.0105 |
| HCM0233 | RUNX1 | chr21 | 36164718 | 36164718 | missense_variant | SNV | G | T | Pro386His | 384 | 10036 | 0.0383 |
| HCM0235 | STAG2 | chrX | 123184974 | 123184974 | missense_variant | SNV | G | A | Gly341Ser | 386 | 152189 | 0.0025 |
| HCM0237 | DNMT3A | chr2 | 25463320 | 25463320 | splice_acceptor_variant | SNV | C | A | c.2174-1G>T | 7454 | 22610 | 0.3297 |
| HCM0240 | TET2 | chr4 | 106156348 | 106156348 | stop_gained | SNV | C | T | Gln417* | 256 | 8458 | 0.0303 |
| HCM0243 | RAD21 | chr8 | 117866652 | 117866652 | missense_variant | SNV | C | A | Glu331Asp | 479 | 21300 | 0.0225 |
| HCM0247 | ASXL1 | chr20 | 31023092 | 31023092 | Frame_shift_Ins | Indel | C | + | Arg860fs | 269 | 14245 | 0.0189 |
| HCM0248 | DNMT3A | chr2 | 25467491 | 25467491 | missense_variant | SNV | C | T | Asp529Asn | 222 | 19679 | 0.0113 |
| HCM0248 | IDH2 | chr15 | 90631934 | 90631934 | missense_variant | SNV | C | T | Arg140Gln | 266 | 24521 | 0.0108 |
| HCM0250 | TET2 | chr4 | 106197380 | 106197380 | missense_variant | SNV | A | G | Lys1905Glu | 88 | 16781 | 0.0052 |

| | | | | | | | | | | | | |
|---|---|---|---|---|---|---|---|---|---|---|---|---|
| HCM0251 | TET2 | chr4 | 106197197 | 106197197 | missense_variant | SNV | G | C | Asp1844His | 5502 | 11700 | 0.4703 |
| HCM0253 | ASXL1 | chr20 | 31022234 | 31022234 | splice_acceptor_variant | SNV | G | A | c.1720-1G>A | 1589 | 16102 | 0.0987 |
| HCM0254 | TP53 | chr17 | 7579358 | 7579358 | missense_variant | SNV | C | A | Arg110Leu | 291 | 30825 | 0.0094 |
| HCM0255 | TET2 | chr4 | 106164073 | 106164073 | missense_variant | SNV | A | G | Ile1195Val | 365 | 2973 | 0.1228 |
| HCM0256 | TET2 | chr4 | 106196309 | 106196309 | stop_gained | SNV | C | T | Gln1548* | 778 | 41257 | 0.0189 |
| HCM0256 | TET2 | chr4 | 106197287 | 106197287 | missense_variant | SNV | G | A | Glu1874Lys | 591 | 9863 | 0.0599 |
| HCM0257 | EZH2 | chr7 | 148508738 | 148508738 | missense_variant | SNV | G | T | Phe642Leu | 538 | 35724 | 0.0151 |
| HCM0260 | DNMT3A | chr2 | 25457159 | 25457159 | missense_variant | SNV | C | G | Ala910Pro | 321 | 12650 | 0.0254 |
| HCM0261 | JAK2 | chr9 | 5073770 | 5073770 | missense_variant | SNV | G | T | Val617Phe | 565 | 23064 | 0.0245 |
| HCM0266 | RUNX1 | chr21 | 36164870 | 36164870 | missense_variant | SNV | C | A | Gln335His | 131 | 482 | 0.2718 |
| HCM0270 | TP53 | chr17 | 7577595 | 7577595 | missense_variant | SNV | C | T | Cys229Tyr | 191 | 23065 | 0.0083 |
| HCM0270 | ASXL1 | chr20 | 31023388 | 31023388 | missense_variant | SNV | C | T | Ser958Leu | 9387 | 20014 | 0.4690 |
| HCM0270 | SMC1A | chrX | 53432728 | 53432728 | missense_variant | SNV | G | T | Thr569Asn | 326 | 22720 | 0.0143 |
| HCM0272 | TET2 | chr4 | 106156729 | 106156729 | stop_gained | SNV | C | T | Arg544* | 1634 | 14421 | 0.1133 |
| HCM0272 | DNMT3A | chr2 | 25457242 | 25457242 | missense_variant | SNV | C | T | Arg882His | 12154 | 55313 | 0.2197 |
| HCM0275 | TET2 | chr4 | 106157163 | 106157163 | Frame_shift_Indel | Indel | T | - | Ser689fs | 529 | 14154 | 0.0374 |
| HCM0275 | FLT3 | chr13 | 28609667 | 28609667 | missense_variant | SNV | C | A | Gly521Val | 421 | 20723 | 0.0203 |
| HCM0277 | ASXL1 | chr20 | 31025110 | 31025110 | missense_variant | SNV | A | G | Lys1532Arg | 159 | 7286 | 0.0218 |
| HCM0283 | PTPN11 | chr12 | 112926945 | 112926945 | missense_variant | SNV | T | C | Ile522Thr | 99 | 17003 | 0.0058 |
| HCM0287 | PPM1D | chr17 | 58740702 | 58740702 | missense_variant | SNV | G | A | Arg536Lys | 6590 | 13130 | 0.5019 |
| HCM0289 | TET2 | chr4 | 106190800 | 106190800 | missense_variant | SNV | C | A | Leu1360Met | 137 | 28475 | 0.0048 |
| HCM0292 | DNMT3A | chr2 | 25468135 | 25468135 | missense_variant | SNV | C | A | Cys514Phe | 265 | 19295 | 0.0137 |
| HCM0294 | DNMT3A | chr2 | 25463560 | 25463560 | missense_variant | SNV | T | G | Ser708Arg | 1412 | 28575 | 0.0494 |
| HCM0296 | PPM1D | chr17 | 58740434 | 58740434 | missense_variant | SNV | G | A | Glu447Lys | 27354 | 55480 | 0.4930 |
| HCM0298 | TET2 | chr4 | 106157376 | 106157377 | Frame_shift_Indel | Indel | TT | - | Phe760fs | 8324 | 23864 | 0.3488 |
| HCM0299 | TET2 | chr4 | 106196927 | 106196927 | missense_variant | SNV | G | C | Gly1754Arg | 32447 | 68400 | 0.4744 |
| HCM0302 | DNMT3A | chr2 | 25469028 | 25469028 | splice_donor_variant | SNV | C | T | c.1429+1G>A | 915 | 11506 | 0.0795 |
| HCM0304 | TP53 | chr17 | 7578466 | 7578466 | missense_variant | SNV | G | A | Thr155Ile | 120 | 7972 | 0.0151 |
| HCM0305 | TET2 | chr4 | 106155502 | 106155502 | stop_gained | SNV | G | T | Glu135* | 86 | 23684 | 0.0036 |
| HCM0305 | CBL | chr11 | 119149000 | 119149000 | missense_variant | SNV | C | A | Ser407Tyr | 280 | 67399 | 0.0042 |
| HCM0310 | TET2 | chr4 | 106158380 | 106158381 | Frame_shift_Indel | Indel | AA | - | Arg1095fs | 735 | 25165 | 0.0292 |

| | | | | | | | | | | | | |
|---|---|---|---|---|---|---|---|---|---|---|---|---|
| HCM0312 | DNMT3A | chr2 | 25467042 | 25467042 | Frame_shift_Indel | Indel | A | - | Asn611fs | 182 | 21927 | 0.0083 |
| HCM0312 | TET2 | chr4 | 106158446 | 106158446 | N | Indel | T | + | Asn1118fs | 149 | 15554 | 0.0096 |
| HCM0316 | CBL | chr11 | 119149218 | 119149218 | splice_acceptor_variant | SNV | A | G | c.1228-2A>G | 105 | 19780 | 0.0053 |
| HCM0316 | IDH1 | chr2 | 209113173 | 209113173 | missense_variant | SNV | T | C | Ile112Val | 4990 | 10790 | 0.4625 |
| HCM0317 | STAG2 | chrX | 123181233 | 123181233 | missense_variant | SNV | G | T | Val233Leu | 486 | 32344 | 0.0150 |
| HCM0322 | DNMT3A | chr2 | 25457176 | 25457176 | missense_variant | SNV | G | A | Pro904Leu | 1720 | 56936 | 0.0302 |
| HCM0323 | DNMT3A | chr2 | 25467073 | 25467073 | stop_gained | SNV | C | T | Trp601* | 236 | 24896 | 0.0095 |
| HCM0327 | ASXL1 | chr20 | 31021496 | 31021496 | missense_variant | SNV | C | T | Arg499Cys | 33688 | 72266 | 0.4662 |
| HCM0328 | TET2 | chr4 | 106196927 | 106196927 | missense_variant | SNV | G | A | Gly1754Ser | 66 | 46494 | 0.0014 |
| HCM0329 | DNMT3A | chr2 | 25470620 | 25470620 | splice_acceptor_variant | SNV | T | C | c.856-2A>G | 62 | 774 | 0.0801 |
| HCM0333 | DNMT3A | chr2 | 25463187 | 25463187 | missense_variant | SNV | A | G | Ile769Thr | 1164 | 10030 | 0.1161 |
| HCM0333 | DNMT3A | chr2 | 25468923 | 25468929 | Frame_shift_Indel | Indel | CACCAGC | - | Leu479fs | 714 | 2782 | 0.2566 |
| HCM0333 | TET2 | chr4 | 106156330 | 106156330 | missense_variant | SNV | C | A | Pro411Thr | 270 | 7737 | 0.0349 |
| HCM0333 | TET2 | chr4 | 106157404 | 106157404 | Frame_shift_Indel | Indel | C | - | Gln769fs | 62 | 13688 | 0.0045 |
| HCM0333 | TET2 | chr4 | 106196666 | 106196666 | missense_variant | SNV | C | A | Leu1667Met | 227 | 10331 | 0.0220 |
| HCM0337 | CEBPA | chr19 | 33792300 | 33792300 | missense_variant | SNV | T | C | Ile341Val | 9524 | 19110 | 0.4984 |
| HCM0339 | KIT | chr4 | 55589807 | 55589807 | missense_variant | SNV | C | A | Ala430Glu | 281 | 11209 | 0.0251 |
| HCM0340 | ZRSR2 | chrX | 15833857 | 15833857 | missense_variant | SNV | G | T | Met205Ile | 268 | 19486 | 0.0138 |
| HCM0342 | TP53 | chr17 | 7578208 | 7578208 | missense_variant | SNV | T | G | His214Pro | 1948 | 63260 | 0.0308 |
| HCM0342 | ASXL1 | chr20 | 31022937 | 31022937 | Frame_shift_Indel | Indel | C | - | Pro808fs | 2046 | 18904 | 0.1082 |
| HCM0348 | ASXL1 | chr20 | 31021206 | 31021206 | missense_variant | SNV | G | A | Arg402Gln | 3744 | 7971 | 0.4697 |
| HCM0353 | PPM1D | chr17 | 58740721 | 58740721 | Frame_shift_Indel | Indel | T | - | Ser543fs | 540 | 34835 | 0.0155 |
| HCM0355 | KRAS | chr12 | 25378647 | 25378647 | missense_variant | SNV | T | G | Lys117Asn | 682 | 6761 | 0.1009 |
| HCM0355 | TET2 | chr4 | 106155911 | 106155911 | stop_gained | SNV | C | G | Ser271* | 840 | 12452 | 0.0675 |
| HCM0355 | TET2 | chr4 | 106157525 | 106157525 | Frame_shift_Ins | Indel | T | + | Gln810fs | 399 | 28380 | 0.0141 |
| HCM0355 | TET2 | chr4 | 106193748 | 106193748 | stop_gained | SNV | C | T | Arg1404* | 1406 | 9859 | 0.1426 |
| HCM0355 | TET2 | chr4 | 106197088 | 106197088 | Frame_shift_Ins | Indel | T | + | Arg1808fs | 5624 | 15358 | 0.3662 |
| HCM0356 | TP53 | chr17 | 7572973 | 7572973 | missense_variant | SNV | C | T | Arg379His | 5456 | 11288 | 0.4833 |
| HCM0356 | TET2 | chr4 | 106193961 | 106193961 | missense_variant | SNV | G | T | Ala1475Ser | 212 | 11458 | 0.0185 |
| HCM0358 | DNMT3A | chr2 | 25462032 | 25462032 | missense_variant | SNV | C | G | Arg792Pro | 278 | 7477 | 0.0372 |
| HCM0360 | IDH2 | chr15 | 90631920 | 90631920 | missense_variant | SNV | C | A | Gly145Trp | 162 | 9492 | 0.0171 |

| | | | | | | | | | | | | |
|---|---|---|---|---|---|---|---|---|---|---|---|---|
| HCM0363 | ASXL1 | chr20 | 31021655 | 31021655 | missense_variant | SNV | A | G | Ile552Val | 8120 | 16479 | 0.4927 |
| HCM0365 | TET2 | chr4 | 106196757 | 106196757 | missense_variant | SNV | G | T | Gly1697Val | 394 | 20005 | 0.0197 |
| HCM0368 | PPM1D | chr17 | 58740727 | 58740727 | Frame_shift_Indel | Indel | C | - | Leu546fs | 174 | 17577 | 0.0099 |
| HCM0368 | STAG2 | chrX | 123220542 | 123220542 | missense_variant | SNV | G | T | Gly1067Trp | 346 | 18825 | 0.0184 |
| HCM0371 | DNMT3A | chr2 | 25469535 | 25469545 | Frame_shift_Indel | Indel | CAGGGCCCAT T | - | Trp409fs | 112 | 20678 | 0.0054 |
| HCM0372 | DNMT3A | chr2 | 25462015 | 25462015 | missense_variant | SNV | G | A | Leu798Phe | 712 | 11710 | 0.0608 |
| HCM0373 | DNMT3A | chr2 | 25463172 | 25463172 | missense_variant | SNV | T | A | Glu774Val | 392 | 71887 | 0.0055 |
| HCM0373 | TET2 | chr4 | 106155328 | 106155328 | Frame_shift_Indel | Indel | G | - | Asp77fs | 144 | 40227 | 0.0036 |
| HCM0377 | ASXL1 | chr20 | 31021206 | 31021206 | missense_variant | SNV | G | A | Arg402Gln | 9847 | 21256 | 0.4633 |
| HCM0381 | TET2 | chr4 | 106157533 | 106157539 | Frame_shift_Indel | Indel | ATAAATC | - | Ile812fs | 185 | 10869 | 0.0170 |
| HCM0382 | TET2 | chr4 | 106164077 | 106164077 | missense_variant | SNV | C | G | Ala1196Gly | 731 | 12442 | 0.0588 |
| HCM0384 | DNMT3A | chr2 | 25463258 | 25463258 | missense_variant | SNV | C | A | Glu745Asp | 296 | 38563 | 0.0077 |
| HCM0384 | RUNX1 | chr21 | 36164646 | 36164646 | missense_variant | SNV | G | A | Ser410Leu | 372 | 34908 | 0.0107 |
| HCM0384 | TET2 | chr4 | 106164769 | 106164769 | stop_gained | SNV | G | A | Val1213Met | 104 | 69386 | 0.0015 |
| HCM0386 | DNMT3A | chr2 | 25457242 | 25457242 | missense_variant | SNV | C | T | Arg882His | 3203 | 63027 | 0.0508 |
| HCM0387 | DNMT3A | chr2 | 25467483 | 25467483 | missense_variant | SNV | G | T | Asp531Glu | 259 | 28699 | 0.0090 |
| HCM0393 | DNMT3A | chr2 | 25459855 | 25459856 | Frame_shift_Indel | Indel | TC | - | Asn810fs | 2875 | 26217 | 0.1097 |
| HCM0393 | FLT3 | chr13 | 28610127 | 28610127 | missense_variant | SNV | C | A | Asp455Tyr | 581 | 25414 | 0.0229 |
| HCM0398 | TET2 | chr4 | 106156280 | 106156280 | missense_variant | SNV | C | A | Ala394Asp | 734 | 41589 | 0.0176 |
| HCM0399 | DNMT3A | chr2 | 25469647 | 25469647 | splice_acceptor_variant | SNV | T | C | c.1123-2A>G | 445 | 26901 | 0.0165 |
| HCM0399 | PPM1D | chr17 | 58740668 | 58740668 | stop_gained | SNV | G | T | Glu525* | 723 | 29409 | 0.0246 |
| HCM0402 | DNMT3A | chr2 | 25469614 | 25469614 | missense_variant | SNV | G | A | Pro385Leu | 427 | 3063 | 0.1394 |
| HCM0404 | TET2 | chr4 | 106196940 | 106196940 | Frame_shift_Indel | Indel | C | - | Ser1758fs | 843 | 95691 | 0.0088 |
| HCM0405 | ASXL1 | chr20 | 31021496 | 31021496 | missense_variant | SNV | C | T | Arg499Cys | 44106 | 93264 | 0.4729 |
| HCM0414 | DNMT3A | chr2 | 25463289 | 25463289 | missense_variant | SNV | T | C | Tyr735Cys | 310 | 13109 | 0.0236 |
| HCM0416 | GNAS | chr20 | 57484407 | 57484407 | missense_variant | SNV | C | A | Asp196Glu | 456 | 73024 | 0.0062 |
| HCM0419 | DNMT3A | chr2 | 25467199 | 25467199 | missense_variant | SNV | C | T | Cys559Tyr | 240 | 18742 | 0.0128 |
| HCM0419 | CBL | chr11 | 119148950 | 119148950 | missense_variant | SNV | T | A | Asp390Glu | 433 | 160667 | 0.0027 |
| HCM0421 | TET2 | chr4 | 106155444 | 106155444 | missense_variant | SNV | C | A | Asp115Glu | 375 | 35157 | 0.0107 |
| HCM0423 | TET2 | chr4 | 106155747 | 106155747 | Frame_shift_Indel | Indel | T | - | Ser217fs | 119 | 18860 | 0.0063 |
| HCM0425 | TET2 | chr4 | 106193934 | 106193934 | stop_gained | SNV | C | T | Gln1466* | 377 | 21959 | 0.0172 |

| | | | | | | | | | | | | |
|---|---|---|---|---|---|---|---|---|---|---|---|---|
| HCM0426 | TET2 | chr4 | 106157356 | 106157356 | Frame_shift_Ins | Indel | A | + | Glu754fs | 4206 | 13605 | 0.3092 |
| HCM0428 | BCOR | chrX | 39914765 | 39914765 | missense_variant | SNV | G | T | Pro1533Thr | 217 | 5773 | 0.0376 |
| HCM0431 | JAK2 | chr9 | 5070010 | 5070010 | missense_variant | SNV | C | A | Asn533Lys | 322 | 23388 | 0.0138 |
| HCM0434 | DNMT3A | chr2 | 25457242 | 25457242 | missense_variant | SNV | C | T | Arg882His | 32819 | 135022 | 0.2431 |
| HCM0435 | ASXL1 | chr20 | 31023843 | 31023843 | missense_variant | SNV | C | A | Gln1110Lys | 362 | 24117 | 0.0150 |
| HCM0436 | ASXL1 | chr20 | 31021590 | 31021590 | missense_variant | SNV | C | A | Ala530Glu | 148 | 15326 | 0.0097 |
| HCM0436 | SMC1A | chrX | 53432233 | 53432233 | stop_gained | SNV | C | A | Glu668* | 344 | 11657 | 0.0295 |
| HCM0437 | DNMT3A | chr2 | 25467432 | 25467432 | missense_variant | SNV | C | A | Met548Ile | 330 | 28245 | 0.0117 |
| HCM0438 | TET2 | chr4 | 106157275 | 106157275 | missense_variant | SNV | C | A | Gln726Lys | 575 | 43955 | 0.0131 |
| HCM0439 | DNMT3A | chr2 | 25470498 | 25470498 | missense_variant | SNV | G | A | Arg326Cys | 413 | 1462 | 0.2825 |
| HCM0440 | PPM1D | chr17 | 58740562 | 58740562 | Frame_shift_Indel | Indel | T | - | Leu490fs | 177 | 16255 | 0.0109 |
| HCM0443 | TET2 | chr4 | 106196771 | 106196771 | missense_variant | SNV | C | A | Gln1702Lys | 333 | 23848 | 0.0140 |
| HCM0446 | DNMT3A | chr2 | 25463181 | 25463181 | missense_variant | SNV | C | T | Arg771Gln | 317 | 17405 | 0.0182 |
| HCM0448 | TET2 | chr4 | 106196941 | 106196941 | Frame_shift_Indel | Indel | A | - | Pro1759fs | 480 | 61099 | 0.0079 |
| HCM0452 | TET2 | chr4 | 106158447 | 106158447 | Frame_shift_Indel | Indel | A | - | Asn1118fs | 37 | 11444 | 0.0032 |
| HCM0455 | FLT3 | chr13 | 28592644 | 28592644 | missense_variant | SNV | C | A | Arg834Leu | 455 | 65649 | 0.0069 |
| HCM0455 | CEBPA | chr19 | 33792387 | 33792387 | missense_variant | SNV | G | T | Gln312Lys | 348 | 17404 | 0.0200 |
| HCM0458 | PPM1D | chr17 | 58740669 | 58740669 | Frame_shift_Indel | Indel | A | - | Ile526fs | 196 | 13496 | 0.0145 |
| HCM0459 | DNMT3A | chr2 | 25463291 | 25463291 | missense_variant | SNV | G | C | Phe734Leu | 95 | 9352 | 0.0102 |
| HCM0462 | TET2 | chr4 | 106190798 | 106190798 | missense_variant | SNV | G | A | Arg1359His | 8197 | 19356 | 0.4235 |
| HCM0469 | DNMT3A | chr2 | 25463287 | 25463287 | missense_variant | SNV | G | A | Arg736Cys | 301 | 32805 | 0.0092 |
| HCM0471 | DNMT3A | chr2 | 25463289 | 25463289 | missense_variant | SNV | T | C | Tyr735Cys | 279 | 35023 | 0.0080 |
| HCM0471 | TET2 | chr4 | 106156384 | 106156384 | missense_variant | SNV | G | A | Gly429Arg | 20601 | 41195 | 0.5001 |
| HCM0478 | DNMT3A | chr2 | 25463287 | 25463287 | missense_variant | SNV | G | A | Arg736Cys | 800 | 30177 | 0.0265 |
| HCM0478 | DNMT3A | chr2 | 25469548 | 25469548 | missense_variant | SNV | A | G | Ile407Thr | 798 | 81432 | 0.0098 |
| HCM0478 | TET2 | chr4 | 106180817 | 106180817 | missense_variant | SNV | G | A | Gly1282Asp | 344 | 31627 | 0.0109 |
| HCM0480 | IDH1 | chr2 | 209113112 | 209113112 | missense_variant | SNV | C | A | Arg132Leu | 234 | 16640 | 0.0141 |
| HCM0484 | PTPN11 | chr12 | 112915523 | 112915523 | missense_variant | SNV | A | G | Asn308Asp | 15932 | 30893 | 0.5157 |
| HCM0486 | RUNX1 | chr21 | 36253003 | 36253003 | missense_variant | SNV | G | T | Ala120Asp | 688 | 88539 | 0.0078 |
| HCM0486 | PPM1D | chr17 | 58740506 | 58740506 | Frame_shift_Indel | Indel | C | - | Pro471fs | 2100 | 71118 | 0.0295 |
| HCM0487 | DNMT3A | chr2 | 25467408 | 25467408 | splice_donor_variant | SNV | C | T | c.1667+1G>A | 287 | 35063 | 0.0082 |

| | | | | | | | | | | | | |
|---|---|---|---|---|---|---|---|---|---|---|---|---|
| HCM0489 | TET2 | chr4 | 106196927 | 106196927 | missense_variant | SNV | G | C | Gly1754Arg | 59929 | 120081 | 0.4991 |
| HCM0492 | DNMT3A | chr2 | 25463289 | 25463289 | missense_variant | SNV | T | C | Tyr735Cys | 312 | 13626 | 0.0229 |
| HCM0495 | ASXL1 | chr20 | 31024748 | 31024748 | stop_gained | SNV | G | A | Trp1411* | 573 | 28050 | 0.0204 |
| HCM0496 | TET2 | chr4 | 106162523 | 106162523 | missense_variant | SNV | C | A | Pro1146His | 361 | 31079 | 0.0116 |
| HCM0499 | DNMT3A | chr2 | 25462020 | 25462020 | missense_variant | SNV | C | T | Gly796Asp | 1405 | 9441 | 0.1488 |
| HCM0499 | GNAS | chr20 | 57484424 | 57484429 | Frame_shift_Indel | Indel | TCCTGA | - | Val202fs | 116 | 34259 | 0.0034 |
| HCM0501 | TET2 | chr4 | 106155904 | 106155904 | missense_variant | SNV | C | A | His269Asn | 775 | 28981 | 0.0267 |
| HCM0504 | TET2 | chr4 | 106157244 | 106157244 | missense_variant | SNV | C | A | Asn715Lys | 340 | 46761 | 0.0073 |
| HCM0505 | DNMT3A | chr2 | 25469528 | 25469528 | missense_variant | SNV | A | T | Phe414Ile | 142 | 26140 | 0.0054 |
| HCM0518 | DNMT3A | chr2 | 25463184 | 25463184 | stop_gained | SNV | G | T | Ser770* | 496 | 20187 | 0.0246 |
| HCM0520 | FLT3 | chr13 | 28592612 | 28592612 | missense_variant | SNV | T | C | Arg845Gly | 275 | 31533 | 0.0087 |
| HCM0521 | DNMT3A | chr2 | 25469520 | 25469523 | Frame_shift_Indel | Indel | AGGC | - | Ser417fs | 133 | 24818 | 0.0054 |
| HCM0522 | DNMT3A | chr2 | 25464456 | 25464456 | missense_variant | SNV | T | C | Asp686Gly | 144 | 7977 | 0.0181 |
| HCM0522 | TET2 | chr4 | 106196528 | 106196528 | missense_variant | SNV | C | T | Leu1621Phe | 64 | 28097 | 0.0023 |
| HCM0522 | TET2 | chr4 | 106196705 | 106196705 | stop_gained | SNV | C | T | Gln1680* | 289 | 23211 | 0.0125 |
| HCM0524 | TET2 | chr4 | 106155751 | 106155751 | missense_variant | SNV | G | T | Val218Leu | 26363 | 53400 | 0.4937 |
| HCM0530 | DNMT3A | chr2 | 25462077 | 25462077 | missense_variant | SNV | G | C | Pro777Arg | 348 | 16139 | 0.0216 |
| HCM0531 | ASXL1 | chr20 | 31021109 | 31021109 | missense_variant | SNV | T | A | Ser370Thr | 5897 | 12271 | 0.4806 |
| HCM0532 | TP53 | chr17 | 7578407 | 7578407 | missense_variant | SNV | G | A | Arg175Cys | 10182 | 22193 | 0.4588 |
| HCM0532 | ZRSR2 | chrX | 15838408 | 15838408 | stop_gained | SNV | C | A | Cys302* | 124 | 17112 | 0.0072 |
| HCM0540 | CALR | chr19 | 13054596 | 13054596 | Frame_shift_Indel | Indel | A | - | Lys375fs | 694 | 9292 | 0.0747 |
| HCM0550 | DNMT3A | chr2 | 25457231 | 25457231 | missense_variant | SNV | G | T | Gln886Lys | 425 | 107244 | 0.0040 |
| HCM0551 | STAG2 | chrX | 123220528 | 123220528 | missense_variant | SNV | G | T | Gly1062Val | 466 | 52580 | 0.0089 |
| HCM0552 | CALR | chr19 | 13054575 | 13054576 | Frame_shift_Indel | Indel | AA | - | Lys368fs | 2128 | 5532 | 0.3847 |
| HCM0552 | DNMT3A | chr2 | 25463265 | 25463265 | missense_variant | SNV | G | C | Pro743Arg | 659 | 31006 | 0.0213 |
| HCM0553 | DNMT3A | chr2 | 25463286 | 25463286 | missense_variant | SNV | C | T | Arg736His | 1009 | 49543 | 0.0204 |
| HCM0553 | TET2 | chr4 | 106182979 | 106182979 | Frame_shift_Ins | Indel | C | + | Leu1340fs | 1284 | 61244 | 0.0210 |
| HCM0553 | TET2 | chr4 | 106197476 | 106197476 | missense_variant | SNV | C | A | Pro1937Thr | 391 | 29549 | 0.0132 |
| HCM0555 | SMC1A | chrX | 53432743 | 53432743 | missense_variant | SNV | C | T | Arg564His | 166 | 11426 | 0.0145 |
| HCM0557 | FLT3 | chr13 | 28610153 | 28610153 | missense_variant | SNV | G | A | Ser446Leu | 6881 | 14593 | 0.4715 |
| HCM0557 | TET2 | chr4 | 106196631 | 106196631 | missense_variant | SNV | C | T | Pro1655Leu | 7606 | 16661 | 0.4565 |

| | | | | | | | | | | | | |
|---|---|---|---|---|---|---|---|---|---|---|---|---|
| HCM0558 | DNMT3A | chr2 | 25463568 | 25463568 | missense_variant | SNV | A | G | Ile705Thr | 1403 | 12271 | 0.1143 |
| HCM0558 | TET2 | chr4 | 106158453 | 106158453 | Frame_shift_Indel | Indel | T | - | Val1136_Glu1137delinsLys | 232 | 48543 | 0.0048 |
| HCM0561 | GNAS | chr20 | 57484468 | 57484468 | missense_variant | SNV | G | T | Val217Phe | 586 | 47838 | 0.0122 |
| HCM0564 | GATA2 | chr3 | 128200763 | 128200763 | missense_variant | SNV | A | G | Cys348Arg | 122 | 7530 | 0.0162 |
| HCM0567 | TET2 | chr4 | 106180816 | 106180816 | missense_variant | SNV | G | T | Gly1282Cys | 362 | 22163 | 0.0163 |
| HCM0570 | DNMT3A | chr2 | 25463184 | 25463184 | missense_variant | SNV | G | A | Ser770Leu | 399 | 16442 | 0.0243 |
| HCM0571 | BCOR | chrX | 39911532 | 39911532 | missense_variant | SNV | G | T | Gln1700Lys | 65 | 2018 | 0.0322 |
| HCM0571 | TET2 | chr4 | 106155184 | 106155184 | missense_variant | SNV | C | A | Pro29Thr | 127 | 852 | 0.1491 |
| HCM0572 | RUNX1 | chr21 | 36253007 | 36253007 | missense_variant | SNV | C | T | Val119Met | 100 | 14649 | 0.0068 |
| HCM0574 | DNMT3A | chr2 | 25462068 | 25462068 | missense_variant | SNV | A | G | Ile780Thr | 139 | 7207 | 0.0193 |
| HCM0577 | FLT3 | chr13 | 28609703 | 28609703 | missense_variant | SNV | C | G | Gly509Ala | 9062 | 20089 | 0.4511 |
| HCM0580 | DNMT3A | chr2 | 25467523 | 25467523 | splice_acceptor_variant | SNV | T | C | c.1555-2A>G | 3058 | 15286 | 0.2001 |
| HCM0589 | TET2 | chr4 | 106190876 | 106190876 | Frame_shift_Ins | Indel | T | + | Leu1385fs | 526 | 22331 | 0.0236 |
| HCM0591 | DNMT3A | chr2 | 25463295 | 25463295 | missense_variant | SNV | T | C | Glu733Gly | 566 | 22794 | 0.0248 |
| HCM0595 | DNMT3A | chr2 | 25470003 | 25470003 | missense_variant | SNV | G | T | Leu347Met | 209 | 11870 | 0.0176 |
| HCM0598 | TET2 | chr4 | 106182928 | 106182928 | stop_gained | SNV | G | T | Glu1323* | 18993 | 49337 | 0.3850 |
| HCM0600 | SMC1A | chrX | 53432850 | 53432850 | missense_variant | SNV | C | A | Lys528Asn | 165 | 16076 | 0.0103 |
| HCM0605 | TET2 | chr4 | 106158505 | 106158508 | Frame_shift_Indel | Indel | GTG | - | Val1136_Glu1137delinsLys | 4319 | 50831 | 0.0850 |
| HCM0609 | CEBPA | chr19 | 33793072 | 33793072 | missense_variant | SNV | C | A | Gln83His | 69 | 494 | 0.1397 |
| HCM0609 | NRAS | chr1 | 115258703 | 115258703 | missense_variant | SNV | G | T | His27Asn | 146 | 2148 | 0.0680 |
| HCM0611 | STAG2 | chrX | 123181341 | 123181341 | stop_gained | SNV | C | T | Gln269* | 137 | 58920 | 0.0023 |
| HCM0612 | DNMT3A | chr2 | 25463308 | 25463308 | missense_variant | SNV | G | A | Arg729Trp | 136 | 9733 | 0.0140 |
| HCM0613 | DNMT3A | chr2 | 25464450 | 25464450 | missense_variant | SNV | C | T | Arg688His | 1347 | 7994 | 0.1685 |
| HCM0613 | ASXL1 | chr20 | 31023641 | 31023641 | Frame_shift_Indel | Indel | A | - | Gly645fs | 1010 | 21951 | 0.0460 |
| HCM0616 | RUNX1 | chr21 | 36171648 | 36171648 | missense_variant | SNV | C | T | Arg306His | 524 | 19493 | 0.0269 |
| HCM0616 | RUNX1 | chr21 | 36206769 | 36206769 | missense_variant | SNV | T | G | Asn248Thr | 3327 | 18955 | 0.1755 |
| HCM0617 | IDH2 | chr15 | 90631935 | 90631935 | missense_variant | SNV | G | A | Arg140Trp | 170 | 7001 | 0.0243 |
| HCM0623 | TP53 | chr17 | 7572988 | 7572988 | missense_variant | SNV | C | A | Gly374Val | 253 | 33380 | 0.0076 |
| HCM0623 | ASXL1 | chr20 | 31021251 | 31021251 | missense_variant | SNV | G | T | Arg417Leu | 231 | 38387 | 0.0060 |
| HCM0627 | DNMT3A | chr2 | 25462075 | 25462075 | missense_variant | SNV | C | T | Val778Met | 1563 | 12452 | 0.1255 |

| | | | | | | | | | | | | |
|---|---|---|---|---|---|---|---|---|---|---|---|---|
| HCM0630 | DNMT3A | chr2 | 25463298 | 25463298 | missense_variant | SNV | A | G | Phe732Ser | 1173 | 13182 | 0.0890 |
| HCM0638 | DNMT3A | chr2 | 25463298 | 25463300 | Frame_shift_Indel | Indel | AAG | - | Phe732fs | 212 | 6143 | 0.0345 |
| HCM0639 | DNMT3A | chr2 | 25469533 | 25469533 | missense_variant | SNV SNV | C | A | Gly412Val | 335 | 17653 | 0.0190 |
| HCM0640 | EZH2 | chr7 | 148516771 | 148516771 | missense_variant | SNV | C | A | Ala306Ser | 233 | 7419 | 0.0314 |
| HCM0641 | TET2 | chr4 | 106190797 | 106190797 | missense_variant | SNV | C | T | Arg1359Cys | 975 | 13774 | 0.0708 |
| HCM0645 | PPM1D | chr17 | 58740956 | 58740956 | 3_prime_UTR_variant | Indel | G | + | n.58740956_58740957insG | 5925 | 11792 | 0.5025 |
| HCM0646 | TET2 | chr4 | 106196491 | 106196491 | stop_gained | SNV | T | G | Tyr1608* | 1211 | 30641 | 0.0395 |
| HCM0656 | TET2 | chr4 | 106197285 | 106197285 | missense_variant | SNV | T | C | Ile1873Thr | 199 | 6776 | 0.0294 |
| HCM0657 | ASXL1 | chr20 | 31021218 | 31021218 | missense_variant | SNV | G | T | Gly406Val | 187 | 7927 | 0.0236 |
| HCM0671 | TET2 | chr4 | 106164911 | 106164911 | missense_variant | SNV | A | G | Asn1260Ser | 4674 | 10590 | 0.4414 |
| HCM0671 | GATA2 | chr3 | 128202759 | 128202759 | missense_variant | SNV | G | T | Leu321Ile | 175 | 1066 | 0.1642 |
| HCM0675 | EZH2 | chr7 | 148506185 | 148506185 | missense_variant | SNV | C | T | Glu725Lys | 385 | 27120 | 0.0142 |
| HCM0675 | EZH2 | chr7 | 148511158 | 148511158 | missense_variant | SNV | C | A | Val582Phe | 252 | 14596 | 0.0173 |
| HCM0680 | TET2 | chr4 | 106158380 | 106158380 | missense_variant | SNV | A | G | Lys1094Arg | 78 | 17592 | 0.0044 |
| HCM0681 | EZH2 | chr7 | 148526830 | 148526830 | missense_variant | SNV | G | T | His158Gln | 410 | 21422 | 0.0191 |
| HCM0687 | CALR | chr19 | 13054560 | 13054560 | stop_gained | SNV | G | T | Glu363* | 273 | 9980 | 0.0274 |
| HCM0689 | TET2 | chr4 | 106193782 | 106193782 | missense_variant | SNV | T | G | Leu1415Arg | 287 | 12368 | 0.0232 |
| HCM0691 | GNAS | chr20 | 57484488 | 57484488 | non_coding_transcript_exon_variant | Indel | C | + | c.659+10del | 9106 | 22176 | 0.4106 |
| HCM0700 | ZRSR2 | chrX | 15836757 | 15836757 | missense_variant | SNV | G | T | Gln273His | 198 | 11462 | 0.0173 |
| HCM0703 | DNMT3A | chr2 | 25463179 | 25463179 | missense_variant | SNV | A | G | Phe772Leu | 2874 | 8353 | 0.3441 |
| HCM0704 | SMC3 | chr10 | 112356244 | 112356249 | Frame_shift_Indel | Indel | AGAGA | - | Ala685fs | 96 | 9597 | 0.0100 |
| HCM0706 | DNMT3A | chr2 | 25464567 | 25464567 | missense_variant | SNV | A | T | Val649Glu | 174 | 5101 | 0.0341 |
| HCM0706 | ASXL1 | chr20 | 31022851 | 31022851 | missense_variant | SNV | C | T | Pro779Leu | 9649 | 20060 | 0.4810 |
| HCM0710 | KRAS | chr12 | 25398228 | 25398228 | stop_gained | SNV | C | A | Glu31* | 209 | 9033 | 0.0231 |
| HCM0712 | SRSF2 | chr17 | 74732959 | 74732959 | missense_variant | SNV | G | A | Pro95Leu | 52 | 1278 | 0.0407 |
| HCM0712 | TET2 | chr4 | 106157257 | 106157257 | stop_gained | SNV | C | T | Gln720* | 191 | 14990 | 0.0127 |
| HCM0712 | TET2 | chr4 | 106157879 | 106157879 | Frame_shift_Indel | Indel | T | - | Pro929fs | 262 | 7886 | 0.0332 |
| HCM0712 | TET2 | chr4 | 106180850 | 106180850 | Frame_shift_Ins | Indel | T | + | Met1293fs | 1176 | 28309 | 0.0415 |
| HCM0712 | TET2 | chr4 | 106180851 | 106180851 | missense_variant | SNV | G | C | Met1293Ile | 1205 | 28307 | 0.0426 |
| HCM0712 | TET2 | chr4 | 106182986 | 106182986 | missense_variant | SNV | C | G | Pro1342Arg | 537 | 17561 | 0.0306 |

| | | | | | | | | | | | | |
|---|---|---|---|---|---|---|---|---|---|---|---|---|
| HCM0712 | TET2 | chr4 | 106182995 | 106182995 | missense_variant | SNV | A | G | Tyr1345Cys | 531 | 17555 | 0.0302 |
| HCM0712 | TET2 | chr4 | 106196424 | 106196424 | stop_gained | SNV | C | A | Ser1586* | 1062 | 11762 | 0.0903 |
| HCM0712 | TET2 | chr4 | 106196758 | 106196758 | Frame_shift_Indel | Indel | A | - | Asn1698fs | 289 | 16617 | 0.0174 |
| HCM0713 | BCOR | chrX | 39933164 | 39933164 | missense_variant | SNV | C | T | Gly479Arg | 3530 | 4777 | 0.7390 |
| HCM0716 | RUNX1 | chr21 | 36164889 | 36164889 | missense_variant | SNV | G | A | Ala329Val | 150 | 8043 | 0.0186 |
| HCM0718 | CBL | chr11 | 119148974 | 119148974 | missense_variant | SNV | C | G | His398Gln | 289 | 44874 | 0.0064 |
| HCM0719 | ASXL1 | chr20 | 31022441 | 31022441 | Frame_shift_Ins | Indel | A | + | Gly643fs | 2780 | 18025 | 0.1542 |
| HCM0721 | CEBPA | chr19 | 33792309 | 33792309 | missense_variant | SNV | G | T | Leu338Met | 444 | 54989 | 0.0081 |
| HCM0722 | DNMT3A | chr2 | 25459832 | 25459832 | missense_variant | SNV | C | A | Glu817Asp | 298 | 61442 | 0.0049 |
| HCM0722 | DNMT3A | chr2 | 25464554 | 25464554 | missense_variant | SNV | C | A | Leu653Phe | 403 | 49955 | 0.0081 |
| HCM0722 | FLT3 | chr13 | 28592705 | 28592705 | missense_variant | SNV | C | G | Ala814Pro | 332 | 95110 | 0.0035 |
| HCM0725 | TET2 | chr4 | 106157218 | 106157218 | missense_variant | SNV | G | T | Ala707Ser | 24417 | 49524 | 0.4930 |
| HCM0729 | CALR | chr19 | 13054650 | 13054658 | Frame_shift_Indel | Indel | GAGGATGAG | - | Glu393fs | 17994 | 36468 | 0.4934 |
| HCM0730 | ASXL1 | chr20 | 31023237 | 31023237 | missense_variant | SNV | G | T | Val908Leu | 370 | 20783 | 0.0178 |
| HCM0737 | U2AF1 | chr21 | 44514770 | 44514770 | missense_variant | SNV | C | A | Glu159Asp | 390 | 13135 | 0.0297 |
| HCM0737 | TET2 | chr4 | 106182947 | 106182948 | Frame_shift_Indel | Indel | TG | - | Ser1330fs | 320 | 53235 | 0.0060 |
| HCM0742 | TET2 | chr4 | 106164929 | 106164929 | missense_variant | SNV | A | G | Asn1266Ser | 913 | 20016 | 0.0456 |
| HCM0745 | TP53 | chr17 | 7573998 | 7573998 | missense_variant | SNV | C | A | Glu343Asp | 309 | 14402 | 0.0215 |
| HCM0745 | ZRSR2 | chrX | 15833805 | 15833805 | stop_gained | SNV | C | A | Ser188* | 462 | 10270 | 0.0450 |
| HCM0745 | ASXL1 | chr20 | 31021550 | 31021550 | missense_variant | SNV | C | A | Gln517Lys | 409 | 26822 | 0.0152 |
| HCM0745 | ASXL1 | chr20 | 31022257 | 31022257 | missense_variant | SNV | C | A | Pro581Gln | 428 | 10761 | 0.0398 |
| HCM0745 | ASXL1 | chr20 | 31024362 | 31024362 | missense_variant | SNV | G | T | Gly1283Cys | 383 | 13429 | 0.0285 |
| HCM0745 | RUNX1 | chr21 | 36206772 | 36206772 | missense_variant | SNV | G | T | Pro247His | 401 | 16322 | 0.0246 |
| HCM0745 | RUNX1 | chr21 | 36206833 | 36206833 | stop_gained | SNV | C | A | Glu227* | 336 | 16666 | 0.0202 |
| HCM0745 | TET2 | chr4 | 106196957 | 106196957 | missense_variant | SNV | C | A | His1764Asn | 1180 | 47943 | |
| HCM0746 | DNMT3A | chr2 | 25467083 | 25467083 | stop_gained | SNV | G | A | Arg598* | 2382 | 42651 | 0.0558 |
| HCM0746 | DNMT3A | chr2 | 25469978 | 25469978 | Frame_shift_Ins | Indel | T | + | His355fs | 187 | 15496 | 0.0121 |
| HCM0748 | SMC1A | chrX | 53432425 | 53432425 | missense_variant | SNV | C | A | Lys637Asn | 266 | 17180 | 0.0155 |
| HCM0750 | DNMT3A | chr2 | 25464487 | 25464487 | missense_variant | SNV | G | A | Arg676Trp | 360 | 23131 | 0.0156 |
| HCM0753 | DNMT3A | chr2 | 25467059 | 25467059 | stop_gained | SNV | G | A | Gln606* | 210 | 28078 | 0.0075 |
| HCM0753 | DNMT3A | chr2 | 25469079 | 25469079 | missense_variant | SNV | C | T | Ser460Asn | 288 | 12555 | 0.0229 |

| | | | | | | | | | | | | |
|---|---|---|---|---|---|---|---|---|---|---|---|---|
| HCM0753 | TET2 | chr4 | 106155973 | 106155973 | missense_variant | SNV | G | T | Val292Leu | 228 | 16594 | 0.0137 |
| HCM0754 | DNMT3A | chr2 | 25469634 | 25469634 | missense_variant | SNV | G | T | Ser378Arg | 533 | 53169 | 0.0100 |
| HCM0757 | CEBPA | chr19 | 33792345 | 33792345 | missense_variant | SNV | T | C | Lys326Glu | 13786 | 30152 | 0.4572 |
| HCM0758 | TP53 | chr17 | 7576911 | 7576911 | missense_variant | SNV | G | C | Thr312Ser | 9014 | 19414 | 0.4643 |
| HCM0762 | DNMT3A | chr2 | 25464441 | 25464441 | missense_variant | SNV | G | T | Thr691Lys | 314 | 9572 | 0.0328 |
| HCM0763 | DNMT3A | chr2 | 25469641 | 25469641 | missense_variant | SNV | G | T | Ala376Asp | 2215 | 79625 | 0.0278 |
| HCM0766 | DNMT3A | chr2 | 25463212 | 25463212 | Frame_shift_Indel | Indel | T | - | Met761fs | 258 | 37990 | 0.0068 |
| HCM0769 | ASXL1 | chr20 | 31024408 | 31024408 | missense_variant | SNV | C | T | Thr1298Ile | 13661 | 28313 | 0.4825 |
| HCM0769 | SMC1A | chrX | 53432559 | 53432559 | missense_variant | SNV | G | T | Leu593Ile | 135 | 11905 | 0.0113 |
| HCM0773 | ASXL1 | chr20 | 31022821 | 31022821 | missense_variant | SNV | C | T | Thr769Ile | 111 | 36412 | 0.0030 |
| HCM0776 | DNMT3A | chr2 | 25463264 | 25463267 | Frame_shift_Indel | Indel | GGGC | - | Pro743del | 338 | 51085 | 0.0066 |
| HCM0782 | BCOR | chrX | 39911617 | 39911617 | missense_variant | SNV | C | A | Lys1671Asn | 373 | 29300 | 0.0127 |
| HCM0782 | SMC3 | chr10 | 112350873 | 112350873 | missense_variant | SNV | G | T | Ala599Ser | 319 | 42608 | 0.0075 |
| HCM0785 | DNMT3A | chr2 | 25463587 | 25463587 | missense_variant | SNV | C | A | Gly699Cys | 237 | 10772 | 0.0220 |
| HCM0786 | PPM1D | chr17 | 58740368 | 58740368 | Frame_shift_Ins | Indel | G | + | Asp425fs | 34 | 10704 | 0.0032 |
| HCM0790 | PTPN11 | chr12 | 112926882 | 112926882 | missense_variant | SNV | G | A | Arg501Lys | 3382 | 8101 | 0.4175 |
| HCM0791 | DNMT3A | chr2 | 25467101 | 25467101 | Frame_shift_Ins | Indel | A | + | Tyr592fs | 1622 | 30081 | 0.0539 |
| HCM0793 | DNMT3A | chr2 | 25457216 | 25457216 | missense_variant | SNV | G | A | Arg891Trp | 2091 | 85390 | 0.0245 |
| HCM0793 | SMC3 | chr10 | 112356246 | 112356246 | missense_variant | SNV | C | A | Ala685fs | 351 | 28746 | 0.0122 |
| HCM0794 | TET2 | chr4 | 106157200 | 106157200 | stop_gained | SNV | C | T | Gln701* | 150 | 17837 | 0.0084 |
| HCM0800 | ASXL1 | chr20 | 31022440 | 31022440 | Frame_shift_Ins | Indel | G | + | Ala215fs | 5734 | 35613 | 0.1610 |
| HCM0800 | TET2 | chr4 | 106156120 | 106156120 | missense_variant | SNV | C | A | Gln341Lys | 243 | 16889 | 0.0144 |
| HCM0800 | TET2 | chr4 | 106157344 | 106157344 | stop_gained | SNV | C | T | Gln749* | 341 | 22430 | 0.0152 |
| HCM0800 | TET2 | chr4 | 106194068 | 106194068 | Frame_shift_Indel | Indel | G | - | Gln1510fs | 311 | 27007 | 0.0115 |
| HCM0801 | TET2 | chr4 | 106190796 | 106190796 | stop_gained | SNV | C | A | Cys1358* | 637 | 34000 | 0.0187 |

| **Supplementary Table 2.** Clinical phenotype and outcomes according to CH among HCM patients. | | | |
|---|---|---|---|
| | All-CH | | |
| | No CH (N=616) | CH (N=183) | p |
| *Echocardiogram* | | | |
| MLVWT, mean (SD), mm | 17.1±4.3 | 16.8±3.9 | 0.352 |
| LA diameter, mm | 36.7±14.5 | 38.1±12.7 | 0.208 |
| LAVi, ml/m$^2$ | 36.5±22.3 | 39.0±25.1 | 0.194 |
| LV EF, % | 60.1±12.9 | 62.5±8.8 | 0.004 |
| LVOT maximal gradient, mm Hg | 24.1±36.2 | 22.1±34.6 | 0.491 |
| SAM, No. (%) | 290 (47.1) | 85 (46.4) | 0.881 |
| Moderate-severe MR, No. (%) | 20 (3.2) | 9 (4.9) | 0.288 |
| *Cardiac MRI* | | | |
| LV mass, g | 154.5±55.9 | 149.5±54.5 | 0.327 |
| LV mass index, g/m$^2$ | 78.1±24.6 | 75.9±24.6 | 0.329 |
| MLVWT, mm | 18.1±5.1 | 17.7±4.3 | 0.401 |
| LV EF, % | 62.0±7.6 | 62.5±6.7 | 0.511 |
| LGE, No (%) | 465 (75.5) | 128 (70.3) | 0.162 |
| LGE >15%, No (%) | 71 (13.7) | 24 (15.5) | 0.583 |
| LV mass % of LGE[1], % | | | |
| Apical Aneurysm, No (%) | 33 (5.4) | 14 (7.7) | 0.004 |
| Syncope, No (%) | 24 (3.9) | 7 (3.8) | 0.965 |
| ABPR at exercise, No. (%) (290/83) | 93 (32.1) | 34 (41) | 0.132 |
| NVST, No. (%) (490/142) | 157 (32.0) | 51 (35.9) | 0.387 |
| *Outcomes* | | | |
| Stroke, No. (%) | 5 (0.8) | 3 (1.6) | 0.323 |
| Appropriate ICD shock, No. (%) | 3 (0.5) | 0 | 0.344 |
| Cardiac arrest, No. (%) | 1 (0.2) | 0 | 0.585 |
| Death or orthotopic heart transplant, No. (%) | 4 (0.6) | 6 (3.3) | 0.005 |
| MACE, No. (%) | 12 (2.0) | 9 (5.2) | 0.026 |

[1]Quantified LGE>5%.

ABPR, abnormal blood pressure response; EF, ejection fraction; LA, left atrium diameter; LAVi, left atrium volume index; ICD, implantable cardioverter defibrillator; LGE, late gadolinium enhancement; LV, left ventricle; LVOT, left ventricular outflow tract; MACE, major cardiovascular events; MLVWT, maximal left ventricular wall thickness; MR, mitral regurgitation; MRI, magnetic resonance imaging; NSVT, non-sustained ventricular tachycardia; SAM, systolic anterior motion;

| **Supplementary Table 3.** Overall characteristics of the HCM cohort and between those with or without CH in *DNMT3A, TET2* and *ASXL1.* | | | |
|---|---|---|---|
| | No CH (N=616) | CH on *DNMT3A, TET2, ASXL1* (N=136) | P |
| Age, mean (SD), years | 55.3±14.6 | 58.1±14.5 | 0.046 |
| Age at diagnosis, mean (SD), years | 47.4±15.6 | 50.0±16.3 | 0.072 |
| Male sex, No. (%) | 424 (68.8) | 88 (64.7) | 0.305 |
| Body mass index[1], mean (SD), $kg/m^2$ | ± | 29.5±6.7 | 0.358 |
| Hypertension, No. (%) | 233 (37.9) | 66 (48.5) | 0.022 |
| Diabetes, No. (%) | 78 (12.7) | 21 (15.4) | 0.646 |
| Prior/current smoker, No. (%) | 122 (19.9) | 30 (22.1) | 0.567 |
| Coronary artery disease, No. (%) | 61 (9.9) | 10 (7.4) | 0.355 |
| Atrial fibrillation, No. (%) | 107 (17.5) | 27 (19.9) | 0.509 |
| Genetic testing, No. (%) | 554 (89.9) | 119 (87.5) | 0.402 |
| Pathogenic/Likely pathogenic variant, No. (%) | 169 (30.5) | 23 (19.3) | 0.014 |
| *MYH7*, No. (%) | 68 (22.1) | 15 (29.4) | 0.492 |
| *MYBPC3*, No. (%) | 147 (47.9) | 21 (41.2) | |
| Other, No. (%) | 92 (30.0) | 15 (29.4) | |
| Family history of HCM, No. (%) | 194 (31.5) | 39 (28.9) | 0.123 |
| Family history of SCD, No. (%) | 26 (4.2) | 9 (6.7) | 0.035 |
| Implantable cardioverter defibrillator, No. (%) | 89 (14.4) | 15 (11.0) | 0.296 |
| Pacemaker, No. (%) | 6 (1.0) | 5 (3.7) | 0.018 |
| NYHA II-IV, No. (%) | 182 (29.6) | 41 (30.1) | 0.907 |
| Beta-blocker, No. (%) | 321 (52.1) | 74 (54.4) | 0.627 |
| Non-dihydropyridine calcium channel blocker, No. (%) | 77 (12.5) | 30 (22.1) | 0.004 |
| Disopyridine, No (%) | 51 (8.3) | 12 (8.8) | 0.836 |
| Diuretic, No (%) | 59 (9.9) | 19 (14.8) | 0.103 |
| Septal reduction therapy, No. (%) | 53 (8.9) | 17 (12.6) | 0.194 |
| Abbreviations: CH, clonal hematopoiesis; HCM, hypertrophic cardiomyopathy; SCD, sudden cardiac death<br>[1]Body mass index calculated as weigh (kg)/height$^2$(m) | | | |

**Supplementary Table 4.** Clinical phenotype and outcomes in the HCM cohort and between those with or without CH in *DNMT3A, TET2* and *ASXL1.*

| | No CH | CH on *DNMT3A, TET2, ASXL1* | |
|---|---|---|---|
| | N=616 | N=136 | P |
| *Echocardiogram* | | | |
| MLVWT, mean (SD), mm | 17.1±4.3 | 16.8±3.9 | 0.427 |
| LA diameter, mm | 36.7±14.5 | 37.5±13.1 | 0.588 |
| LAVi, ml/m$^2$ | 36.5±22.3 | 36.5±25.8 | 0.995 |
| LV EF, % | 60.1±12.9 | 63.0±8.3 | 0.013 |
| LVOT maximal gradient, mm Hg | 24.1±36.2 | 23.0±35.8 | 0.739 |
| SAM, No. (%) | 290 (47.1) | 64 (47.1) | 0.997 |
| Moderate-severe MR, No. (%) | 20 (3.2) | 7 (5.1) | 0.281 |
| *Cardiac MRI* | | | |
| LV mass, g | 154.1±55.9 | 149.7±55.3 | 0.402 |
| LV mass index, g/m$^2$ | 78.1±24.6 | 75.5±25.4 | 0.316 |
| MLVWT, mm | 18.1±5.1 | 17.6±4.3 | 0.312 |
| LV EF, % | 62.1±7.6 | 62.4±6.5 | 0.629 |
| LGE, No (%) | 465 (75.5) | 96 (71.1) | 0.289 |
| LGE >15%, No (%) | 71 (13.7) | 17 (12.5) | 0.795 |
| LV mass % of LGE[1], % | 8.9±8.7 | 9.2±10.5 | 0.803 |
| Apical Aneurysm, No (%) | 33 (5.4) | 10 (7.3) | 0.374 |
| Syncope, No (%) | 24 (3.9) | 5 (3.7) | 0.904 |
| ABPR at exercise, No. (%) (290/83) | 93 (32.1) | 29 (21.3) | 0.081 |
| NVST, No. (%) (490/142) | 157 (32.0) | 42 (30.8) | 0.268 |
| *Outcomes* | | | |
| Stroke, No. (%) | 5 (0.8) | 2 (1.5) | 0.469 |
| Appropriate ICD shock, No. (%) | 3 (0.5) | 0 | 0.415 |
| Cardiac arrest, No. (%) | 1 (0.2) | 0 | 0.638 |
| Death or orthotopic heart transplant, No. (%) | 4 (0.6) | 6 (4.4) | <0.0001 |
| MACE, No. (%) | 12 (2.0) | 8 (6.2) | 0.009 |

[1]Quantified LGE>5%.

ABPR, abnormal blood pressure response; EF, ejection fraction; LA, left atrium diameter; LAVi, left atrium volume index; ICD, implantable cardioverter defibrillator; LGE, late gadolinium enhancement; LV, left ventricle; LVOT, left ventricular outflow tract; MACE, major cardiovascular events; MLVWT, maximal left ventricular wall thickness; MR, mitral regurgitation; MRI, magnetic resonance imaging; NSVT, non-sustained ventricular tachycardia; SAM, systolic anterior motion;

**Supplementary Table 5.** Overall characteristics of the HCM cohort with germline P/LP sarcomeric variants among those with or without CH .

| | Without germline sarcomeric gene LP/P | | With germline sarcomeric gene LP/P | | All | Without germline sarcomeric gene LP/P<br><br>No CH vs. CH | With germline sarcomeric gene LP/P<br><br>No CH vs. CH |
|---|---|---|---|---|---|---|---|
| | No CH (N=360) | CH (N=120) | No CH (N=169) | CH (N=37) | P | P | P |
| Age, mean (SD), years | 57.8±13.3 | 58.9±13.5 | 50.6±16.1 | 50.5±17.5 | <0.0001 | 0.414 | 0.975 |
| Age at diagnosis, mean (SD), years | 50.6±14.2 | 52.2±14.7 | 40.9±16.3 | 40.0±18.7 | <0.0001 | 0.289 | 0.765 |
| Male sex, No. (%) | 261 (72.5) | 86 (71.7) | 100 (59.2) | 21 (56.8) | 0.004 | 0.860 | 0.787 |
| Body mass index[1], mean (SD), $kg/m^2$ | 30.1±6.1 | 29.5±6.5 | 26.6±8.4 | 27.8±4.8 | <0.0001 | 0.386 | 0.442 |
| Hypertension, No. (%) | 163 (45.3) | 64 (53.3) | 40 (23.8) | 8 (21.6) | <0.0001 | 0.126 | 0.776 |
| Diabetes, No. (%) | 55 (15.3) | 19 (15.8) | 14 (8.3) | 4 (10.8) | 0.332 | 0.710 | 0.630 |
| Prior/current smoker, No. (%) | 83 (23.1) | 22 (18.3) | 25 (14.9) | 11 (29.7) | 0.185 | 0.385 | 0.032 |
| Coronary artery disease, No. (%) | 46 (12.8) | 8 (6.7) | 7 (4.2) | 0 | 0.002 | 0.067 | 0.206 |
| Atrial fibrillation, No. (%) | 54 (15.0) | 20 (16.7) | 27 (16.1) | 8 (21.6) | 0.658 | 0.661 | 0.417 |
| *MYH7*, No. (%) | n/a | n/a | 43 (25.6) | 10 (27.0) | | | |
| *MYBPC3*, No. (%) | n/a | n/a | 106 (63.1) | 21 (56.8) | n/a | n/a | 0.664 |
| Other, No. (%) | n/a | n/a | 19 (11.3) | 6 (16.2) | | | |
| Family history of HCM, No. (%) | 59 (16.4) | 28 (23.3) | 107 (63.3) | 21 (56.8) | <0.0001 | 0.060 | 0.565 |
| Family history of SCD, No. (%) | 9 (2.5) | 7 (5.8) | 15 (8.9) | 3 (8.1) | 0.032 | 0.121 | 0.559 |
| Implantable cardioverter defibrillator, No. (%) | 47 (13.1) | 9 (7.5) | 38 (22.5) | 8 (21.6) | 0.004 | 0.101 | 0.909 |
| Pacemaker, No. (%) | 4 (1.0) | 3 (2.5) | 0 | 2 (5.4) | 0.032 | 0.155 | 0.002 |
| NYHA II-IV, No. (%) | 115 (31.9) | 31 (25.8) | 47 (28.0) | 15 (40.5) | 0.271 | 0.208 | 0.132 |

| | | | | | | | |
|---|---|---|---|---|---|---|---|
| Beta-blocker, No. (%) | 203 (56.4) | 66 (55.0) | 74 (43.8) | 20 (54.1) | 0.026 | 0.791 | 0.256 |
| Non-dihydropyridine calcium channel blocker, No. (%) | 58 (16.1) | 31 (25.8) | 12 (7.1) | 5 (13.5) | <0.0001 | 0.018 | 0.199 |
| Disopyridine, No (%) | 36 (10.0) | 12 (10.0) | 6 (3.6) | 2 (5.4) | 0.077 | 1.00 | 0.597 |
| Diuretic, No (%) | 43 (12.3) | 15 (13.0) | 9 (5.5) | 2 (5.6) | 0.044 | 0.831 | 0.981 |
| Septal reduction therapy, No. (%) | 27 (7.5) | 16 (13.3) | 18 (11.1) | 4 (10.8) | 0.285 | 0.055 | 0.958 |

Abbreviations: CH, clonal hematopoiesis; HCM, hypertrophic cardiomyopathy; P/LP, pathogenic/likely pathogenic; SCD, sudden cardiac death
[1]Body mass index calculated as weigh (kg)/height$^2$(m)

**Supplementary Table 6.** Overall characteristics of the HCM cohort with germline P/LP sarcomeric variants and between those with or without CH in *DNMT3A, TET2* and *ASXL1.*

| | No CH (N=169) | CH on *DNMT3A, TET2 AND ASXL1* (N=23) | P |
|---|---|---|---|
| Age, mean (SD), years | 50.7±16.1 | 49.9±16.7 | 0.831 |
| Age at diagnosis, mean (SD), years | 41.1±16.3 | 37.5±17.7 | 0.353 |
| Male sex, No. (%) | 100 (59.2) | 11 (47.8) | 0.301 |
| Body mass index[1], mean (SD), kg/m$^2$ | 26.6±8.4 | 28.3±5.7 | 0.381 |
| Hypertension, No. (%) | 40 (23.8) | 4 (17.4) | 0.493 |
| Diabetes, No. (%) | 14 (8.3) | 1 (4.3) | 0.505 |
| Prior/current smoker, No. (%) | 25 (14.9) | 7 (30.4) | 0.061 |
| Coronary artery disease, No. (%) | 7 (4.2) | 0 | 0.319 |
| Atrial fibrillation, No. (%) | 27 (16.1) | 4 (17.4) | 0.872 |
| *MYH7*, No. (%) | 43 (25.6) | 7 (30.4) | 0.829 |
| *MYBPC3*, No. (%) | 106 (63.1) | 13 (56.5) | |
| Other, No. (%) | 19 (11.3) | 3 (13.0) | |
| Family history of HCM, No. (%) | 107 (63.3) | 12 (52.2) | 0.470 |
| Family history of SCD, No. (%) | 15 (8.9) | 3 (13.0) | 0.589 |
| Implantable cardioverter defibrillator, No. (%) | 38 (22.5) | 6 (26.1) | 0.700 |
| Pacemaker, No. (%) | 0 | 0 | -- |
| NYHA II-IV, No. (%) | 47 (28.0) | 10 (43.5) | 0.128 |
| Beta-blocker, No. (%) | 74 (43.8) | 13 (56.5) | 0.250 |
| Non-dihydropyridine calcium channel blocker, No. (%) | 12 (7.1) | 2 (8.7) | 0.783 |
| Disopyridine, No (%) | 6 (3.6) | 0 | 0.359 |
| Diuretic, No (%) | 9 (5.5) | 1 (4.5) | 0.859 |
| Septal reduction therapy, No. (%) | 18 (11.1) | 1 (4.3) | 0.317 |

Abbreviations: CH, clonal hematopoiesis; HCM, hypertrophic cardiomyopathy; P/LP, pathogenic/likely pathogenic; SCD, sudden cardiac death

[1]Body mass index calculated as weigh (kg)/height$^2$(m)

| **Supplementary Table 7.** Overall characteristics of the HCM cohort and between those with or without CH after propensity score matching. | | | |
|---|---|---|---|
| | No CH (N=514) | CH (N=183) | P |
| Age, mean (SD), years | 56.2±14.7 | 57.2±14.8 | 0.443 |
| Age at diagnosis, mean (SD), years | 48.4±15.9 | 49.6±16.3 | 0.398 |
| Male sex, No. (%) | 344 (66.9) | 123 (67.2) | 0.943 |
| Body mass index[1], mean (SD), kg/m$^2$ | 29.0±6.9 | 29.2±6.2 | 0.654 |
| Hypertension, No. (%) | 217 (42.3) | 82 (44.8) | 0.556 |
| Diabetes, No. (%) | 72 (14.1) | 26 (14.2) | 0.961 |
| Prior/current smoker, No. (%) | 96 (18.6) | 36 (19.6) | 0.564 |
| Coronary artery disease, No. (%) | 56 (10.8) | 12 (6.5) | 0.092 |
| Atrial fibrillation, No. (%) | 74 (14.3) | 30 (16.3) | 0.521 |
| Genetic testing, No. (%) | 464 (90.2) | 158 (86.3) | 0.140 |
| Pathogenic/Likely pathogenic variant, No. (%) | 143 (27.8) | 37 (20.2) | 0.181 |
| *MYH7*, No. (%) | 88 (17.1) | 21 (11.4) | 0.758 |
| *MYBPC3*, No. (%) | 37 (7.1) | 10 (5.4) | |
| Other, No. (%) | 17 (3.3) | 6 (3.2) | |
| Family history of HCM, No. (%) | 194 (37.7) | 57 (31.1) | 0.084 |
| Family history of SCD, No. (%) | 26 (5.0) | 11 (6.0) | 0.048 |
| Implantable cardioverter defibrillator, No. (%) | 89 (17.3) | 22 (12.0) | 0.405 |
| Pacemaker, No. (%) | 6 (1.1) | 7 (3.8) | 0.007 |
| NYHA II-IV, No. (%) | 182 (35.4) | 53 (28.9) | 0.859 |
| Beta-blocker, No. (%) | 321 (62.4) | 100 (54.6) | 0.547 |
| Non-dihydropyridine calcium channel blocker, No. (%) | 77 (14.9) | 39 (21.3) | 0.003 |
| Disopyridine, No (%) | 51 (9.9) | 16 (8.7) | 0.842 |
| Diuretic, No (%) | 59 (11.4) | 22 (12.0) | 0.303 |
| Septal reduction therapy, No. (%) | 53 (10.3) | 23 (12.5) | 0.130 |
| Abbreviations: CH, clonal hematopoiesis; HCM, hypertrophic cardiomyopathy; SCD, sudden cardiac death<br>[1]Body mass index calculated as weigh (kg)/height$^2$(m) | | | |

| **Supplementary Table 8.** Clinical phenotype and outcomes in the HCM cohort and between those with or without CH after propensity matching. | | | |
|---|---|---|---|
| | No CH N=514 | CH N=183 | P |
| *Echocardiogram* | | | |
| MLVWT, mean (SD), mm | 16.8±4.1 | 16.8±3.9 | 0.948 |
| LA diameter, mm | 36.6±14.5 | 38.1±12.8 | 0.194 |
| LAVi, ml/m$^2$ | 36.4±22.9 | 39.0±25.2 | 0.202 |
| LV EF, % | 60.2±12.9 | 62.5±8.9 | 0.027 |
| LVOT maximal gradient, mm Hg | 23.4±35.1 | 22.1±34.6 | 0.662 |
| SAM, No. (%) | 237 (46.1) | 85 (46.7) | 0.890 |
| Moderate-severe MR, No. (%) | 17 (3.3) | 9 (4.9) | 0.317 |
| *Cardiac MRI* | | | |
| LV mass, g | 151.9±55.0 | 149.2±54.6 | 0.603 |
| LV mass index, g/m$^2$ | 76.7±24.2 | 75.7±24.6 | 0.687 |
| MLVWT, mm | 17.7±4.5 | 17.7±4.3 | 0.949 |
| LV EF, % | 62.1±7.5 | 62.5±6.7 | 0.596 |
| LGE, No (%) | 379 (73.7) | 127 (69.3) | 0.353 |
| LGE >15%, No (%) | 54 (10.5) | 24 (13.1) | 0.364 |
| LV mass % of LGE[1], % | 8.5±8.4 | 9.5±10.1 | 0.347 |
| Apical Aneurysm, No (%) | 18 (3.5) | 8 (4.3) | 0.135 |
| Syncope, No (%) | 19 (3.7) | 7 (3.8) | 0.927 |
| ABPR at exercise, No. (%) (290/83) | 82 (15.9) | 34 (18.5) | 0.169 |
| NVST, No. (%) (490/142) | 150 (29.1) | 51 (27.8) | 0.841 |
| *Outcomes* | | | |
| Stroke, No. (%) | 4 (0.8) | 3 (1.6) | 0.201 |
| Appropriate ICD shock, No. (%) | 3 (0.6) | 0 | 0.302 |
| Cardiac arrest, No. (%) | 1 (0.2) | 0 | 0.552 |
| Death or orthotopic heart transplant, No. (%) | 4 (0.8) | 6 (3.3) | 0.014 |
| MACE, No. (%) | 11 (2.2) | 9 (5.2) | 0.049 |

[1]Quantified LGE>5%.

ABPR, abnormal blood pressure response; EF, ejection fraction; LA, left atrium diameter; LAVi, left atrium volume index; ICD, implantable cardioverter defibrillator; LGE, late gadolinium enhancement; LV, left ventricle; LVOT, left ventricular outflow tract; MACE, major cardiovascular events; MLVWT, maximal left ventricular wall thickness; MR, mitral regurgitation; MRI, magnetic resonance imaging; NSVT, non-sustained ventricular tachycardia; SAM, systolic anterior motion;

| **Supplementary Table 9.** Cytokines and chemokines expression among HCM patients with sarcomeric mutations according to CH-associated gene mutations. | | | |
|---|---|---|---|
| | No CH (N=169) | CH (N=37) | P |
| CCL21 | 275.5 (195.3-365.3) | 386.2 (244.7-538.2) | 0.36 |
| BNP | 19.4 (14.8-30.8) | 21.1 (17.1-28.5) | 0.631 |
| Troponin I | 298.9 (77.1-557.7) | 444.0 (260.6-675.8) | 0.008 |
| sCD40L | 120.0 (70.4-172.8) | 124.7 (79.4-347.8) | 0.280 |
| EGF | 12.6 (5.7-21.0) | 7.9 (5.3-32.4) | 0.826 |
| CCL-11 | 22.8 (17.5-29.6) | 28.9 (20.5-43.5) | 0.079 |
| FGF-2 | 39.9 (27.1-93.7) | 49.7 (30.3-93.7) | 0.114 |
| FLT3 | 5.4 (3.2-7.1) | 6.4 (3.5-11.1) | 0.079 |
| CX3CL1 | 103.3 (67.9-186.2) | 98.9 (67.9-148.8) | 0.956 |
| G-CSF | 43.0 (27.0-71.3) | 56.6 (31.0-206.9) | 0.088 |
| GM-CSF | 30.8 (6.9-10.7.2) | 51.7 (5.6-268.5) | 0.135 |
| CXCL1 | 4.4 (2.1-11.1) | 6.1 (2.1-10.2) | 0.386 |
| IFNα2 | 47.4 (30.8-72.5) | 63.5 (35.8-108.4) | 0.706 |
| IFNγ | 2.2 (1.1-6.5) | 1.8 (1.2-22.1) | 0.430 |
| IL-1α | 22.1 (16.1-46.0) | 23.5 (15.1-47.0) | 0.261 |
| IL-1β | 7.2 (3.8-13.0) | 6.9 (2.9-19.1) | 0.335 |
| IL-1RA | 4.8 (3.0-8.4) | 6.1 (4.4-7.8) | 0.037 |
| IL-2 | 0.9 (0.4-2.7) | 0.8 (0.5-3.2) | 0.255 |
| IL-3 | 0.8 (0.4-1.4) | 1.0 (0.6-1.6) | 0.501 |
| IL-4 | 0.5 (0.2-1.3) | 0.6 (0.1-2.5) | 0.115 |
| IL-5 | 3.9 (2.6-6.2) | 4.4 (2.1-12.0) | 0.072 |
| IL-6 | 1.5 (0.9-3.2) | 2.2 (1.5-4.5) | 0.028 |
| IL-7 | 1.0 (0.5-2.3) | 2.1 (0.4-3.7) | 0.421 |
| IL-8 | 2.5 (1.6-3.5) | 2.3 (1.7-4.0) | 0.122 |
| IL-9 | 6.0 (2.4-11.4) | 5.3 (3.0-15.1) | 0.256 |
| IL-10 | 2.3 (1.0-4.2) | 2.8 (1.2-4.5) | 0.359 |
| IL-12p40 | 63.3 (41.3-117.8) | 63.3 (34.7-298.3) | 0.292 |
| IL-12p70 | 7.9 (5.0-14.2) | 7.2 (5.3-17.5) | 0.478 |
| IL-13 | 75.0 (44.1-173-7) | 70.1 (44.9-262.6) | 0.951 |
| IL-15 | 4.2 (2.9-6.4) | 5.0 (2.6-9.3) | 0.161 |
| IL-17A | 3.7 (1.5-6.1) | 3.2 (1.7-12.9) | 0.148 |
| IL-17E/IL-25 | 716.5 (387.9-1065.6) | 9506 (425.4-3198.5) | 0.321 |
| IL-17F | 21.9 (12.2-57.7) | 45.2 (22.8-524.9) | 0.006 |
| IL-18 | 129.1 (83.0-216.2) | 164.2 (109.7-211.3) | 0.942 |
| IL-22 | 125.0 (70.9-227.3) | 162.8 (97.6-316.8) | 0.247 |
| IL-27 | 1269.2 (933.0-1653.0) | 1120.1 (744.3-2003.5) | 0.767 |
| CXCL10 | 145.2 (90.9-228.3) | 123.4 (91.8-230.1) | 0.508 |
| CCL2 | 196.6 (136.9-240.7) | 184.4 (153.1-279.3) | 0.733 |
| CCL7 | 13.4 (9.3-23.9) | 14.0 (8.2-27.5) | 0.670 |
| CSCF1 | 30.7 (13.6-62.5) | 28.7 (21.9-35.7) | 0.340 |
| CXCL12 | 700.3 (469-875) | 610.6 (418.2-742.3) | 0.795 |
| CXCL9 | 1263.8 (961.9-1802.0) | 1463.3 (1050.1-2201.0) | 0.566 |
| CCL3 | 32.2 (25.0-41.8) | 37.3 (27.4-47.8) | 0.156 |
| CCL4 | 29.3 (19.3-42.1) | 34.6 (20.7-54.6) | 0.318 |
| PDGF-AA | 145.5 (86.2-253.4) | 121.0 (83.1-263.8) | 0.661 |
| PDGF-AA/BB | 3538.2 (2336.5-5313.0) | 3453.6 (2158.3-4720.3) | 0.414 |
| CCL5 | 2372.9 (1883.4-3024.6) | 2133.9 (155.0-3361.9) | 0.769 |
| TGF-α | 6.8 (3.0-16.8) | 13.1 (7.7-488.5) | 0.005 |
| TNF-α | 41.1 (30.3-82.2) | 34.9 (20.2-81.3) | 0.444 |
| TNF-β | 17.7 (11.3-36.1) | 18.1 (10.5-29.7) | 0.892 |

| | | | |
|---|---|---|---|
| VEGF-A | 9.3 (4.6-19.8) | 12.5 (4.8-28.9) | 0.149 |
| CXCL13 | 52.0 (38.1-78.7) | 45.8 (29.9-100.0) | 0.982 |
| CCL27 | 1263.5 (1014.2-1528.0) | 1273.5 (1053.8-1588.7) | 0.367 |
| CXCL5 | 149.0 (82.3-265.3) | 159.7 (90.0-375.2) | 0.823 |
| CCL24 | 389.8 (201.5-653.2) | 463.7 (225.8-615.4) | 0.147 |
| CCL26 | 19.4 (16.2-37.0) | 26.3 (18.1-98.2) | 0.112 |
| CCL1 | 2.3 (1.6-3.5) | 3.4 (1.9-5.1) | 0.002 |
| IL-16 | 356.8 (1.4-1122.1) | 275.9 (2.4-976.6) | 0.752 |
| IL-20 | 907.2 (579.6-1301.3) | 734.9 (461.3-1462.1) | 0.830 |
| IL-21 | 7.0 (2.8-27.4) | 15.4 (2.4-56.4) | 0.218 |
| IL-23 | 812.2 (55.-5730.4) | 4774.9 (419.9-22830.9) | 0.164 |
| IL-28A | 222.5 (2.4-2157.2) | 271.9 (2.4-1747.4) | 0.846 |
| IL-33 | 16.4 (4.7-80.1) | 53.7 (6.9-606.0) | 0.065 |
| LIF | 12.7 (5.3-29.3) | 28.5 (6.1-217.5) | 0.129 |
| CCL8 | 31.9 (25.3-42.3) | 42.8 (27.4-57.8) | 0.036 |
| CCL13 | 47.1 (32.0-106.0) | 83.5 (48.3-250.6) | 0.070 |
| CCL15 | 4362.8 (2957.1-5808.7) | 4251.2 (3210.6-6151.4) | 0.769 |
| SCF | 38.0 (22.0-82.8) | 47.0 (19.0-198.7) | 0.112 |
| CXCL12 | 4633.6 (3433.6-6161.1) | 4858.5 (1933.4-7937.8) | 0.962 |
| CCL17 | 20.3 (14.2-28.1) | 25.2 (13.8-41.5) | 0.047 |
| TPO | 216.0 (72.7-738.6) | 568.1 (161.7-7052.9) | 0.293 |
| TRAIL | 66.0 (49.8-106.0) | 81.7 (39.5-105.2) | 0.103 |
| TSLP | 5.9 (1.7-34.3) | 14.4 (1.5-352.9) | 0.725 |
